# Exploring the application of behaviour change technique taxonomies in childhood obesity prevention interventions: A systematic scoping review

**DOI:** 10.1101/2022.01.25.22269797

**Authors:** Debapriya Chakraborty, Bronwyn A Bailey, Anna Lene Seidler, Serene Yoong, Kylie E Hunter, Rebecca K Hodder, Angela C Webster, Brittany J Johnson

## Abstract

Behaviour change technique (BCT) taxonomies provide one approach to unpack the complexity of childhood obesity prevention interventions. This scoping review sought to examine how BCT taxonomies have been applied to understand childhood obesity prevention interventions targeting children aged 12 years or under and/or their caregivers. A systematic search was conducted in Medline, Embase, PsycINFO, Cochrane Central Register of Controlled Trials, Cochrane Database of Systematic Reviews, CINAHL and PROSPERO. Eligible studies included any study design that applied a BCT taxonomy and evaluated behavioural childhood obesity prevention interventions targeting children aged 12 years or under and/or their parents or caregivers. Sixty-three records, describing 54 discrete studies were included; 32 applied a BCT taxonomy prospectively (i.e., to design interventions) and 23 retrospectively (i.e., to assess interventions), 1 study did both. There was substantial variation in the methods used to apply BCT taxonomies and to report BCT-related methods and results. There was a paucity of detail reported in how BCTs were selected in studies applying BCT taxonomies prospectively. Our review provides important insight into the application of BCT taxonomies in childhood obesity prevention and several ongoing challenges, pointing to the need for best practice reporting guidance.

## INTRODUCTION

The prevalence of obesity globally has reached epidemic proportions. Previously a problem of high-income countries, recently childhood overweight and obesity rates have also been rising in low and middle-income countries[1]. Worldwide in 2019, 38 million children under 5 years and 340 million children aged between 5 and 19 years were experiencing obesity or overweight[1]. Early prevention of obesity in childhood is a health priority as children affected by obesity are much more likely to be affected by obesity as adults, leading to a higher susceptibility to developing chronic diseases at younger ages[1].

Parents and caregivers, and children themselves, should be supported to prevent obesity by developing healthy energy balance behaviours, such as positive infant feeding, optimal dietary intake, sufficient activity levels and sleep. Interventions for childhood prevention of obesity have been studied extensively, with results showing mixed effectiveness[2-4]. As the determinants of obesity are complex and varied, no one single approach is likely to prevent childhood obesity[5]. Interventions to prevent childhood obesity will need to incorporate a variety of approaches across different settings, making them very complex[5]. This complexity presents challenges to upscaling or reproducing interventions, as well as to understanding important components driving behaviour change[6].

Behaviour Change Technique (BCT^1^) taxonomies provide a method to characterise interventions informed by behavioural theory principles[7]. Behaviour Change Techniques are defined as the smallest identifiable, reproducible components of an intervention that can cause a change in behaviour[7]. Taxonomies have been developed to provide a standardised process to describe behaviour change content across interventions[8]. A systematic approach to classifying the contents of complex behaviour change interventions via a taxonomy enables the identification of potentially effective components in both primary studies and systematic reviews[9]. Out of the different taxonomies available[10-12], the BCT Taxonomy v1 (*BCTTv1*) is the most comprehensive, is multi-disciplinary, and has been foundational to the progression of behaviour change science[13]. By nature, application of taxonomies has a level of subjectivity, meaning there may be variability in how they are utilised. While training is often available, application may differ based on researcher expertise, target populations, type of interventions, and level of detail in intervention descriptions. Previous studies have assessed the impact of training on BCT coding and reporting of interventions targeting adult populations, finding both positive and mixed impacts[13, 14]. A previous scoping review examined the methods used by researchers to determine effectiveness of BCTs in changing health-related behaviour, however, this review did not examine the methods used to select or code BCTs[8]. Consistency in the selection and application of BCTs (in prospective studies) as well as extraction and coding (in retrospective studies) is needed to 1) increase the utility of BCTs to better describe the active components of child obesity prevention intervention; 2) allow for higher quality evidence-synthesis and 3) provide opportunity to understand the impact of specific BCTs on improving obesity-related outcomes. Overall, knowledge is limited on the methods of BCT taxonomies application in research relating to childhood obesity prevention. Critique of the use and methods of applying BCT taxonomies is needed to give confidence in evidence syntheses determining effective BCTs for this population. In addition, insight into such methods can identify best practice approaches and provide guidance for childhood obesity prevention researchers in future studies.

This review sought to answer the question: *How have BCT taxonomies been applied to understand childhood obesity prevention interventions targeting children aged ≤12 years, and/or their caregivers?* The focus of this review was to explore the methods used by researchers in applying such taxonomies in both prospective (i.e., when designing or adapting an intervention) and retrospective (i.e., post-hoc use in reviews or secondary analyses) applications. Specifically, this review sought to: 1) categorise the design and key characteristics of studies that have applied a BCT taxonomy, including the timing of BCT taxonomy application (i.e., prospectively or retrospectively) and frequency of application over time; and 2) describe the methods used in applying BCT taxonomies.

## METHODS AND MATERIALS

This review followed a systematic approach, with a protocol prospectively registered on Open Science Framework Registries (https://osf.io/83fgw). Reporting of this review follows the Preferred Reporting Items for Systematic reviews and Meta-Analyses extension for Scoping Reviews (PRISMA-ScR) checklist[15] (Supplementary File 1).

### Eligibility criteria

Studies were eligible if they included: a) a population of children aged ≤12 years at baseline (including prenatal period) and/or their parents or caregivers; and b) applied a BCT taxonomy in relation to child obesity prevention intervention(s), such as Abraham and Michie’s (2008) BCT taxonomy, ‘Coventry, Aberdeen & London – Refined’ (CALO-RE) taxonomy, BCTTv1[7, 11, 12, 16] (i.e. the phenomenon of interest). Studies that included a broader population were only eligible if greater than or equal to 80% of participants were within the targeted child age group. Childhood obesity prevention interventions were defined as those aiming to change obesity-related behaviour(s), such as those relating to infant feeding (e.g., breastfeeding, formula feeding, introduction of solids), diet, movement (e.g., physical activity, sedentary behaviour), and sleep.

Any study design involving the primary or secondary analysis of childhood obesity prevention interventions were eligible. Prospective BCT taxonomy application could include study designs that report interventions, including individual or cluster randomised controlled trials, interrupted time series, quasi-randomised trials, pre-post studies, and intervention design studies with a qualitative component. Types of publications could include protocols, intervention development or primary outcome papers. Retrospective BCT taxonomy application could include reviews of interventions, secondary analyses or critiques of interventions.

Studies were excluded if they only involved children with overweight or obesity (since the focus of this study was on prevention not treatment), severe illness or chronic conditions that impacted their weight status or related behaviour, if they focused on prevention of stunting or underweight, or if the active intervention did not extend beyond the prenatal period.

### Search

Systematic searches were conducted in February 2021 in Medline (Ovid), Embase (Ovid), PsycINFO (Ovid), Cochrane Central Register of Controlled Trials (CENTRAL), Cochrane Database of Systematic Reviews, and Cumulative Index to Nursing and Allied Health Literature (CINAHL; EBSCO). Unpublished studies were searched for via the International Prospective Register of Systematic Reviews (PROSPERO) and indirectly via CENTRAL, which includes registration records from the WHO International Clinical Trials Registry Portal and ClinicalTrials.gov. Targeted searches of clinical trials registers were beyond the time and resource constraints for this scoping review[17]. The search strategy included keywords relating to the population of interest (e.g. ‘child’, ‘infant’, ‘parent’), and to the application of a BCT taxonomy (e.g. ‘behaviour change technique’), in the context of child obesity prevention (e.g. ‘obesity’, ‘overweight’). There were no restrictions on country, language, publication dates, or full text availability. The full search strategy is included in Supplementary File 2. Reference lists of included studies were examined to identify additional eligible records.

### Selection of sources of evidence

All titles and abstracts were screened in Covidence systematic review software (Veritas Health Innovation Melbourne Australia) by two independent reviewers. There was 92% agreement between reviewers for title and abstract screening, and conflicts were resolved by a third reviewer. When uncertainty remained, the reference was included for full text screening. The full texts were each screened by two independent reviewers, and differences were resolved by discussion (DC, BAB, BJJ). At the full text stage there was 90% agreement between reviewers. Forward-backward citation searches were undertaken to identify additional publications describing included studies, for example to identify a final publication where only a PROSPERO registration or protocol publication were retrieved in the database search. Where multiple included records described the same study, all available records were used to complete data charting.

### Data charting process and data items

For each included study, data were extracted by one reviewer using Microsoft Excel (Microsoft Corporation, Redmond, WA), with a random 50% sample independently verified by a second reviewer. Any disagreements that arose after data verification by a second reviewer were resolved by the senior reviewer. The data extraction tool was developed based on researcher expertise and pilot tested on a small sample of studies, then revised and finalised for data extraction. Data extraction included general study characteristics, methods and reporting of BCT taxonomy application (**Table 1**). Authors of one study were contacted to request supplementary files which could not be located online. Critical appraisal of studies was not performed as the focus of this scoping review was on the methods implemented, rather than evaluating results and effectiveness of interventions.

**Table 1:** Summary of data extraction tool items

| Category | Items |
| --- | --- |
| General study characteristics | *For all study designs:*<br>• authors<br>• geographic location<br>• publication date<br>• study design<br>• study population<br>*For retrospective studies:*<br>• number of individual studies included |
| Methods and reporting of BCT taxonomy application | *For all study designs:*<br>• population target of the BCTs<br>• target behaviour/s<br>• name of BCT taxonomy<br>• BCT training undertaken by authors<br>• BCT numbers and labels<br>• total number of BCTs used/coded<br>• details of each unique BCT identified<br>*For prospective studies:*<br>• process of how BCTs were selected<br>*For retrospective studies:*<br>• type of information used to code BCTs such as published materials (i.e., intervention descriptions in publications) and/or unpublished materials (e.g., facilitator handbooks, participant materials)<br>• number of coders<br>• independence of coders<br>• process for managing discrepancies in coding (if applicable) |
|  | • how the BCT results were synthesised |

### Synthesis of results

Characteristics of included studies and applied BCT methods were narratively synthesised and presented descriptively in summary tables, to allow comparisons of key characteristics, frequency of use over time, and the BCT methods applied. Findings were summarised separately for prospective and retrospective applications of a BCT taxonomy, as while there were some common methods and reporting criteria there were several different methodological aspects related to each of these categories of studies. To explore commonly used BCTs across studies, unique reported BCTs were mapped against the 93 BCTs in the BCTTv1[7], for both prospective and retrospective applications.

## RESULTS

### Selection of sources of evidence

A total of 6725 records were identified, with 63 records selected, describing 54 discrete studies (**Figure 1**; Supplementary File 3). Of the 54 studies, results were available for 49, while 4 were registrations only and 1 was a pre-print of a protocol. Thirty-two studies were classified as prospective applications of a BCT taxonomy, most commonly of randomised controlled trial (RCT) study design (n=16/32; including cluster-randomised and pilot RCTs), followed by studies describing intervention development (n=7/32). Twenty-three studies were classified as retrospectively applying a BCT taxonomy. The most common retrospective study design was systematic reviews (n=15/23); other retrospective study designs were multi-method study (n=2/23), secondary analysis (n=1/23), methodology study (n=1/23), exploratory study with content analysis (n=1/23), systematic assessment of mobile applications (n=1/23), intervention development (n=1/23), and scoping review (n=1/23). The median number of primary studies assessed in the studies classified as retrospectively applying a BCT taxonomy was 16 and ranged from one [18, 19] to 64[20]. One of the included studies applied a BCT taxonomy both prospectively and retrospectively as it involved adaption of an intervention to a new context[19].

**Figure 1:**
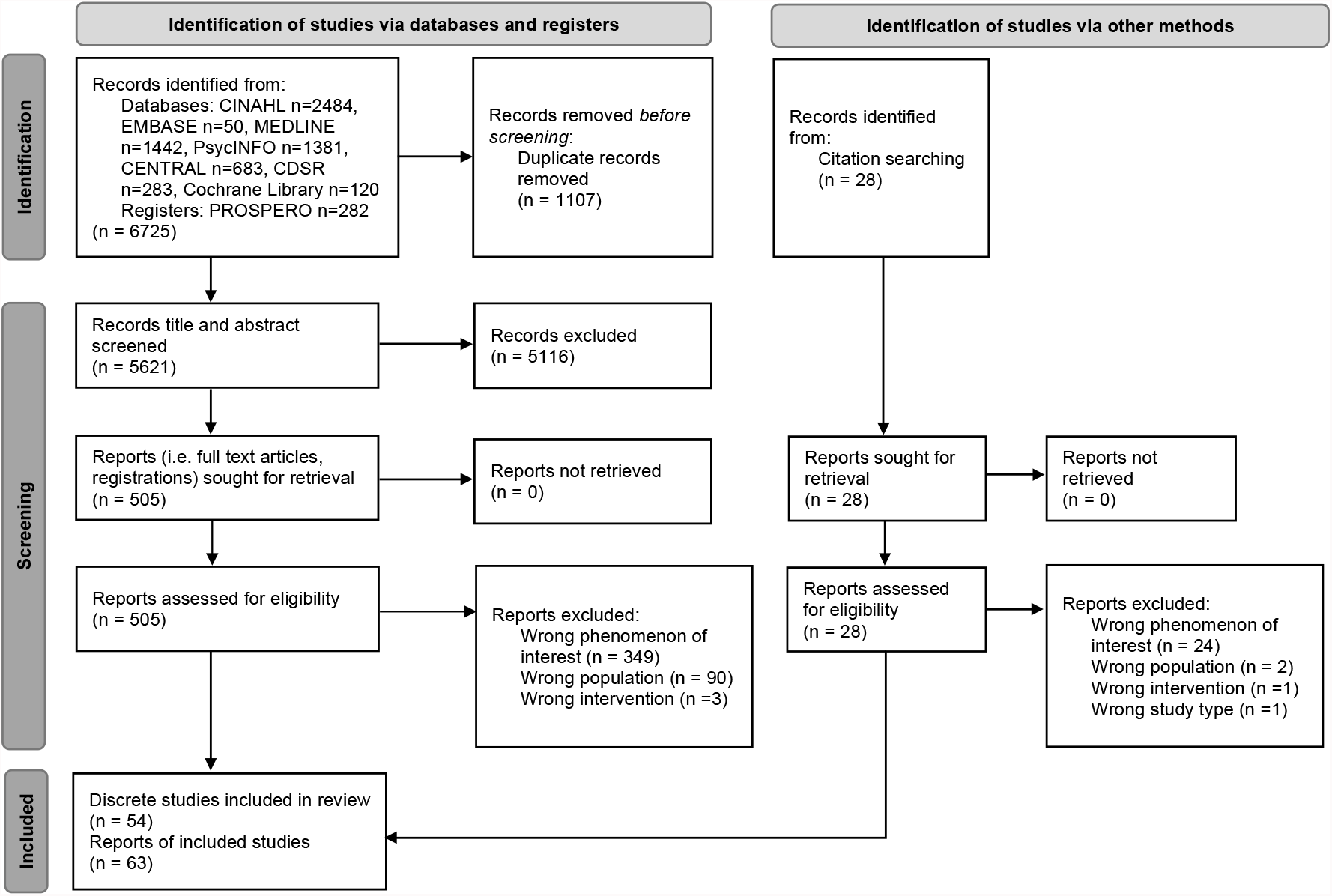
Preferred Reporting Items for Systematic Reviews and Meta-Analyses (PRISMA) flow diagram of search results and selection of re cords

### Characteristics and methods applied in prospective use of BCT taxonomies

**Table 2** presents a summary of prospective BCT taxonomy applications in childhood obesity prevention. Of the 32 studies identified that prospectively used the BCT taxonomy, the BCTs were most frequently targeting parents and families (n=22) or children (n=13). Physical activity (n=28) and dietary intake (n=23) were the most targeted behaviours, followed by infant feeding (n=7), while sleep health (n=3) was the least targeted behaviour.

**Table 2:**
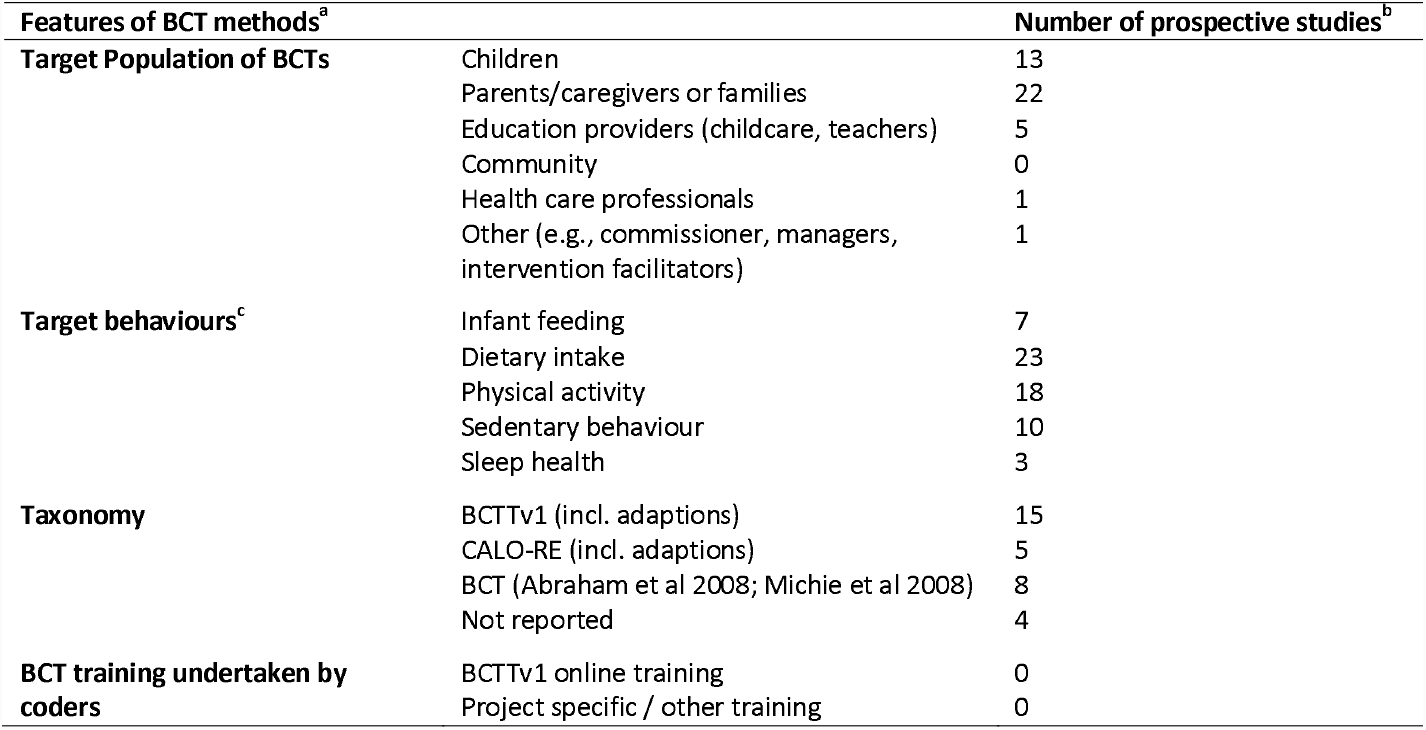
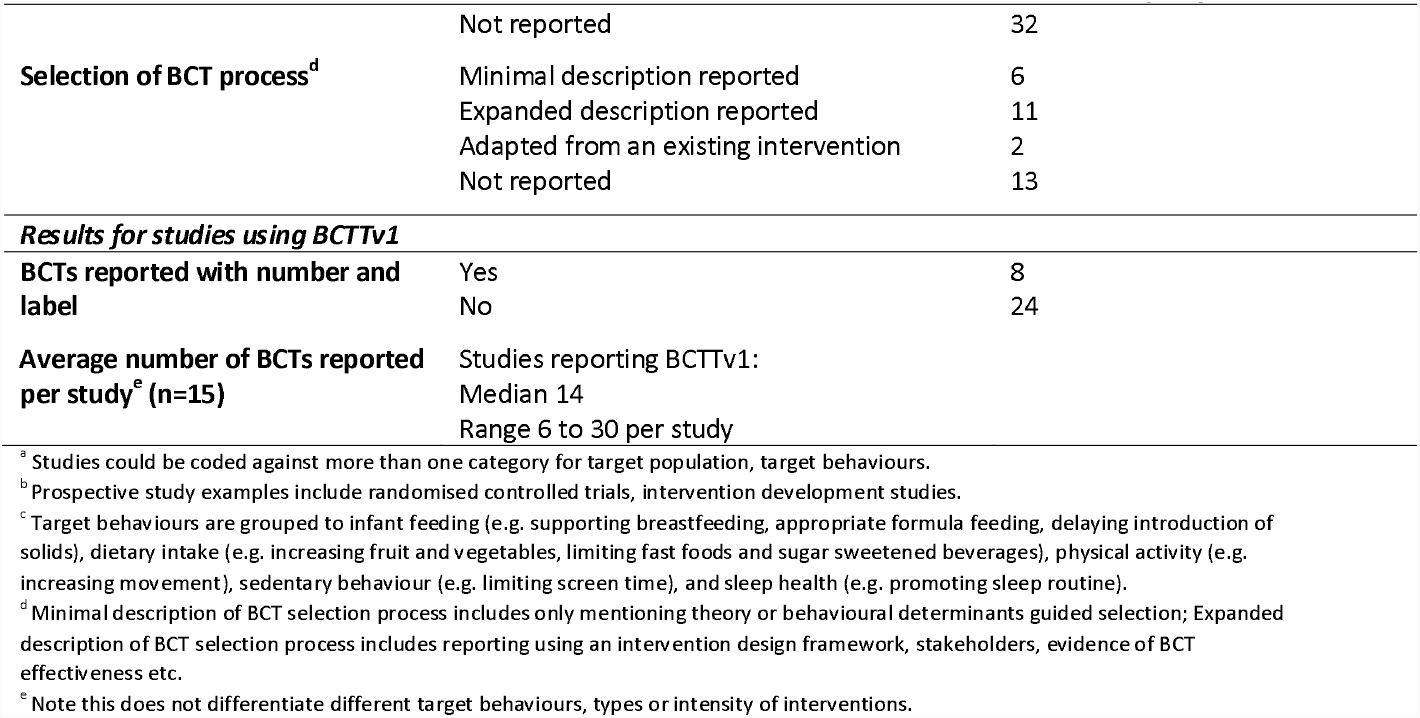
Summary of prospective BCT taxonomy applications in childhood obesity prevention (n=32)

The BCTTv1[7] was the most frequently used taxonomy to code and develop intervention content (n=15/32), followed by the BCT taxonomy[11] (n=8/32). The process to select BCTs was often not reported (n=13/32) or reported with minimal description (n=6/32). No study reported whether researchers had completed BCT taxonomy training. Approximately one third of studies (n=11/32) reported using an intervention design framework, stakeholders and/or evidence of BCT effectiveness to guide BCT selection. Of the studies that utilised the BCTTv1 taxonomy, about half reported the BCTs with the numbers and labels from the taxonomy (n=8/15).

The majority of studies were published by researchers in the UK (n=9/32), USA (n=8/32), and Australia (n=7/32). The remainder in Ireland (n=2/32), New Zealand (n=2/32), China (n=1/32), Germany (n=1/32), Mexico (n=1/32), and South Africa (n=1/32). More than half of the studies were published in the past 5 years (n=18/32; 2021 n=2, 2020 n=4, 2019 n=5, 2018 n=6, 2017 n=1), with most (n=12/18) using the BCTTv1 taxonomy, which was published in 2013[7]. See Supplementary Table 1 for details by individual prospective study.

Across prospective studies a total of 56 of 93 unique BCTs were identified from BCTTv1, plus an additional 5 techniques developed by study authors (Supplementary Table 2). Of the studies that used the BCTTv1 (n=15), reported number of BCTs ranged from 6[21-24] to 30[25], with a median number of 14. The most frequently identified BCTs (identified in 9 or more studies) were *4*.*1 Instruction on how to perform behaviour* (n=12), *5*.*1 Information about health consequences* (n=11), 1.1 *Goal setting (behaviour)* (n=9), *2*.*3 Self-monitoring of behaviour* (n=9), and *6*.*1 Demonstration of the behaviour* (n=9)[7].

### Characteristics and methods applied in retrospective use of BCT taxonomies

**Table 3** presents a summary of retrospective BCT taxonomy applications in childhood obesity prevention. Of the 23 studies identified that retrospectively used the BCT taxonomy, the BCTs were most frequently targeting parents and families (n=15) or children (n=7). Physical activity (n=15) and dietary intake (n=13) were the most targeted behaviours, followed by infant feeding (n=8), and sleep health (n=6).

**Table 3:** Summary of BCT taxonomy retrospective applications in childhood obesity prevention (n=23)

| Features of BCT methods<sup>a</sup> |  | Number of retrospective studies<sup>b</sup> |
| --- | --- | --- |
| Target Population of BCTs | Children | 7 |
|  | Parents/caregivers or families | 15 |
|  | Education providers (childcare, teachers) | 1 |
|  | Community | 2 |
|  | Health care professionals | 0 |
| Target behaviours<sup>c</sup> | Infant feeding | 8 |
|  | Dietary intake | 13 |
|  | Physical activity | 8 |
|  | Sedentary behaviour | 7 |
|  | Sleep health | 6 |
| Type(s) of intervention materials coded<sup>d</sup> | Published only | 12 |
|  | Unpublished only | 2 |
|  | Published and unpublished | 4 |
|  | App / website / activity tracker | 2 |
|  | Not reported | 3 |
| Taxonomy | BCTTv1 (incl. adaptations) | 18 |
|  | CALO-RE (incl. adaptations) | 2 |
|  | BCT (Abraham et al 2008) | 1 |
|  | Other / Not reported | 2 |
| Number of coders per study | 1<sup>d</sup> | 5 |
|  | 2 | 12 |
|  | 3 | 1 |
|  | 4 | 1 |
|  | Not reported | 4 |
| Independence of coders | Independent | 16 |
|  | Unclear or not reported | 6 |
| Process for managing discrepancies in coding | Discussion between coders | 14 |
|  | Third person / expert consulted<sup>e</sup> | 10 |
| BCT training undertaken by coders | BCTTv1 online training | 10 |
|  | Project specific / other training | 4 |
|  | Not reported | 12 |
| BCTs synthesis procedure<sup>f,g</sup> | Narrative summary | 15 |
|  | Quantitative analysis (e.g., meta-regression) | 6 |
|  | Not reported | 2 |
| **Results for studies using BCTTv1** |  |  |
| BCTs reported with number and label<sup>f</sup> | Yes | 12 |
|  | No | 6 |

| Features of BCT methods<sup>a</sup> | Number of retrospective studies<sup>b</sup> |
| --- | --- |
| **Average number of BCTs reported per primary study<sup>c</sup>** | Studies reporting BCTTv1:<br>Published materials only (n=8)<br>5.6 median (4.3 to 9)<br>Unpublished materials (only or in addition to published) (n=4)<br>17.5 median (13.9 to 45) |

<sup>a</sup> Studies could be coded against more than one category for target population, target behaviours, process for managing discrepancies, BCT training undertaken by coders and BCTs synthesis procedure.
<sup>b</sup> Retrospective studies include systematic reviews, multi-method, secondary analysis, methodology study, exploratory study with content analysis, systematic assessment of applications, intervention development.
<sup>c</sup> Target behaviours are grouped to infant feeding (e.g. supporting breastfeeding, appropriate formula feeding, delaying introduction of solids), dietary intake (e.g. increasing fruit and vegetables, limiting fast foods and sugar sweetened beverages), physical activity (e.g. increasing movement), sedentary behaviour (e.g. limiting screen time), and sleep health (e.g. promoting sleep routine).
<sup>d</sup> All studies with 1 coder double coded a subset (10 to 22% double coded or n=20 double coded) or verified coding by another coder.
<sup>e</sup> This category captures both the primary approach of consulting a third coder, or only if needed following discussion between coders.
<sup>f</sup> For studies with results available (n=18).
<sup>g</sup> Note this does not differentiate between behaviours targeted, types or intensity of interventions.

BCTs were most frequently coded from published materials only (n=12/23), chiefly using the BCTTv1 taxonomy (n=18/23), by two coders (n=12/23) or one primary coder and a proportion cross checked (n=5/23). Coding was predominately independent (n=16/23), with discrepancies resolved through discussion (n=14/23) or by consulting a third person (n=10/23). Approximately half of the studies did not report coder training (n=12/23). Of those that reported coder training, most reported BCTTv1 online training (n=10/11). Of the studies that utilised the BCTTv1 taxonomy, two thirds reported the BCTs with the number and label from the taxonomy (n=12/18), the remainder reported only BCT labels or different phrasing to describe BCTs (e.g. ‘goal setting’, rather than specifying if it referred to BCT *1*.*1 Goal setting* (*behaviour*) or BCT *1*.*3 Goal setting* (*outcome*)). Majority of the BCT results were synthesised and reported as narrative summaries (n=15/23), with few using varied quantitative synthesis approaches. Most retrospective studies were published in the past 5 years (n=22/23; 2021 n=3, 2020 n=7, 2019 n=5, 2018 n=3, 2017 n=4), with most (n=18/22) using the BCTTv1 taxonomy. See Supplementary Table 3 for details by individual retrospective study.

Across retrospective studies a total of 80 of 93 unique BCTs were identified from BCTTv1, as well as 4 unofficial BCTs that authors added to the taxonomy for their study (Supplementary Table 4). Of those studies which used the BCTTv1 (n=15), the median number of BCTs reported per study was 5.6, ranging from 4.3[26] to 9[27] for published reports. The median number of BCTs reported when including unpublished materials was 17.5, ranging from 13.9[18] to 45[28] BCTs. Unpublished materials included items such as manuals for intervention delivery, participant handouts, phone scripts, transcriptions from intervention sessions, and unpublished intervention trial protocols. The number of BCTs identified when coding app / website / activity tracker was a mean of 3.9. Most frequently identified BCTs (utilised in 10 or more studies) were *3*.*1 Social support (unspecified)* (n=15), *1*.*1 Goal setting (behaviour)* (n=14), *1*.*4 Action planning* (n=14), *1*.*2 Problem solving* (n=13), *4*.*1 Instruction on how to perform a behaviour* (n=13), *5*.*1 Information about health consequences* (n=13), *7*.*1 Prompts/cues* (n=13), *2*.*3 Self-monitoring of behaviour* (n=13), *6*.*1 Demonstration of the behaviour* (n=12), *8*.*1 Behavioural practice/rehearsal* (n=12), *12*.*5 Adding objects to the environment* (n=12), *3*.*2 Social support (practical)* (n=11), *12*.*1 Restructuring the physical environment* (n=11), *1*.*5 Review behaviour goal(s)* (n=10), and *2*.*2 Feedback on behaviour* (n=10)[7].

## DISCUSSION

Interventions to change obesity-related behaviours in childhood are extremely complex, and BCT taxonomies provide one approach to unpack this complexity. Prior to this review there was limited literature describing the landscape of research applying BCT taxonomies in child obesity prevention. This review has explored how BCT taxonomies have been applied to understand childhood obesity prevention interventions targeting children aged ≤12 years and/or their caregivers. We found limited detail in the reporting of BCT taxonomy-related methods, particularly in prospective applications of BCT taxonomies, indicating the need for best practice guidelines for reporting such methods. There was large variation in the number of BCTs reported or identified when using only published materials compared with unpublished materials. Finally, we found five BCTs that were commonly used in childhood obesity prevention interventions, regardless of prospective or retrospective BCT application. This review provides a snapshot of the use of BCT taxonomies in childhood obesity prevention and direction to improve applications in this evolving research field.

### Opportunities to improve reporting of the application of BCT taxonomies

Limited details of methods to identify BCTs were reported in both prospective and retrospective applications of BCT taxonomies. Methods details were particularly scarce in prospective applications. Specific details on BCTs reporting are needed to support evidence syntheses and provide opportunity to understand the impact of specific BCTs on improving obesity-related outcomes. For example, intervention protocols only reporting the names of selected BCTs, with no details of the process undertaken to select the BCTs. Selected BCTs were rarely reported with their corresponding number and label from the taxonomy, and in some instances the BCT taxonomy used was not stated. Within studies that retrospectively coded BCTs, approximately half did not report coder training and a third did not report BCTs with their corresponding number and label from a taxonomy. Poor methods reporting has implications for our ability to synthesise BCT effectiveness, particularly when there is uncertainty regarding which BCTs were used (i.e., no number and label). It also reduces confidence in the reliability of BCTs coded from existing interventions (e.g., unclear of training, number of coders and independence, types of materials)[13].

Poor reporting is not unique to BCT taxonomy applications. There have been calls to increase the quality of reporting of behavioural intervention descriptions with the development of the TIDieR checklist and CONSORT-SPI extension[9, 10]. Yet, such guidance does not include reporting of BCT-related processes although an understanding of these components are crucial to unpack the ‘black box’ of intervention core components. We propose similar reporting guidance is needed for retrospective use of BCT taxonomies, potentially as a PRISMA-BCT extension for reporting systematic reviews that code BCTs. In addition, guidance regarding methods for BCT selection and reporting would assist transparency and quality use of taxonomies in prospective applications. Such guidance could supplement coding instructions provided with the BCTTv1 online training for example[29]. Researchers have previously provided methods guidance that researchers undertake BCT taxonomy training[29]. In addition, a previous review of the different methods to determine BCT effectiveness, could be used to inform recommendations for BCT synthesis[8]. Finally, methods aspects examined in our review provide direction for expanding guidance to include processes to select BCTs, transparency in coding procedures and clear results presentation. This will increase confidence in understanding the selection of BCTs in future interventions and reviews. See **Box 1** for preliminary recommendations for reporting BCT taxonomy applications.

#### Box 1

Preliminary recommendations for reporting BCT taxonomy applications in childhood obesity prevention interventions

**Prospective taxonomy applications** (*i*.*e. in the design of interventions*)

- Name the taxonomy and provide the citation
- Detail the researchers selecting BCTs, including any BCT training and prior experience
- Detail the process used to select BCTs in sufficient detail to allow replication, including how each BCT is operationalised in the intervention
- List of all included BCTs, reported with the corresponding number and label from the taxonomy

**Retrospective taxonomy applications** (*i*.*e. assessing interventions*)

- Name the taxonomy and provide the citation
- Detail the researchers coding BCTs, including the number of coders in total, BCT training and prior experience
- Detail the types of materials used and coding process, including, number of coders per trial, independence of coders, consensus processes
- List of all identified BCTs, reported with the corresponding number and label from the taxonomy
- Detail the methods used to synthesis BCT findings

### Variation in the number of BCTs identified with types of materials coded

We observed large variation in the number of BCTs identified in retrospective applications of taxonomies when coding only published materials (i.e., publications), compared with unpublished intervention materials (e.g., participant resources) and those reported in prospective studies. The difference in the average number of BCTs identified per study when coding published versus unpublished materials was approximately 12 BCTs. This suggests coding BCTs from publications alone may underestimate the true range of unique BCTs included in an intervention. Publication of intervention protocols and use of supplementary files to share all unpublished materials can increase the detail of information available for an intervention, to confidently code BCTs. A broad range of intervention types were eligible for the review so this may reflect differences in BCTs in large, multi-component interventions versus discrete brief interventions, this requires further examination. An alternate explanation may be that when coding unpublished intervention materials that BCTs of lower dose or unintended BCTs are able to be identified. Hence coding of unpublished materials may inflate the number of BCT compared with those planned, or additional BCTs may not add to understanding the behaviour change process.

The median number of BCTs reported by trialists in prospective applications (14 BCTs) was similar to that per individual trial when coding unpublished intervention descriptions, which may support the prior hypothesis that BCTs are under reported in published materials. Developing reporting guidance as proposed above would assist researchers to clearly and transparently report the rationale for BCT selection and aid future evidence synthesis by decreasing reliance on coding unpublished materials. However, a larger sample of retrospective studies that have coded unpublished materials is needed to draw firmer conclusions.

### Commonly used BCTs in childhood obesity prevention interventions

A range of unique BCTs were selected prospectively (n=15; 56 of 93) or identified retrospectively (n=15; 80 of 93) in childhood obesity prevention interventions, when applying the BCTTv1[7]. There were several BCTs that were common across study designs, including *1*.*1 Goal setting (behaviour), 2*.*3 Self-monitoring of behaviour, 4*.*1 Instruction on how to perform behaviour, 5*.*1 Information about health consequences*, and *6*.*1 Demonstration of behaviour*. Use of such BCTs may suggest common inclusion of intervention components seeking to change individual’s knowledge and motivation. Such BCTs, with the exception of *6*.*1 Demonstration of behaviour*, have also been commonly reported in systematic reviews of adult obesity prevention or management interventions[30] or interventions targeting dietary intake and/or physical activity in adult populations[31-33]. In addition, the adult literature has commonly identified *2*.*2 Feedback on behaviour* and *3*.*1 Social support (unspecified)*[30-33]. These BCTs were frequently reported in retrospective studies in the current review (BCT 2.2 n=10; BCT 3.1 n=13), but less commonly reported in prospective studies (BCT 2.2 n=3; BCT 3.1 n=5, respectively). There were only 13 BCTs that were not reported in any study in our review, several of which are unlikely applicable for this research area and target population, such as *8*.*5 Overcorrection, 10*.*11 Future punishment, 11*.*1 Pharmacological support*, and *11*.*4 Paradoxical instructions*. There were however 20 unique BCTs used in only 1 or 2 prospective studies and 30 unique BCTs reported in only 1 or 2 retrospective studies. The less frequently used BCTs could be incorporated into existing interventions to increase their impact or in new interventions to evaluate their potential to change behaviour in childhood obesity prevention.

### Strengths and limitations of the review

Our study is the first to assemble and examine the BCT methods used in the prevention of childhood obesity and thus identified specific opportunities to advance the methods around this. The review is strengthened by the systematic search and processes undertaken to identify relevant literature. Similarly, the planned approach was to have two reviewers independently extract data, however, given the number of records identified data extraction was completed by one reviewer with a percentage cross-checked by another reviewer, which is common for scoping reviews and few discrepancies were identified. There were several challenges faced in synthesising the methods used in applying BCT taxonomies in this review, generally because of scarce detail in the methods reported in publications. It is important to consider the findings of this review within the confines of reported intervention detail. For example, in prospective applications training in BCT taxonomies was rarely reported, however researchers may have undertaken such training but not reported this within the intervention publication. There was inherent English language bias despite no search limits being applied for language. We found studies were primarily from the USA, UK and Australia. This potential English language or geography bias may be a result of the taxonomies being developed and published in English; future translations of such taxonomies may support uptake of BCT taxonomies in other regions. A final limitation is that data extraction of retrospective studies did not record the frequency of BCT application in the included intervention trials of each study, but rather the total number of unique BCTs and mean number of BCTs identified across trials.

### Future directions

The current delve into the application of BCT taxonomies in childhood obesity prevention research has revealed several additional research questions that warrant investigation. Firstly, there is a need to develop best practice guidance for BCT taxonomy application and reporting. Such a process should involve expert consensus involving researchers with expertise in BCT taxonomies and childhood obesity prevention intervention research. Variation in the number of BCTs identified when using different types of intervention descriptions leads to numerous further methods investigations that were out of scope of the current review. It would be helpful to explore the underrepresentation of BCTs in publications or unintended BCTs when examining unpublished materials. If such future research highlights the need to code unpublished materials, guidance will be needed for which types of materials to be prioritised and establish a process for researchers to share open-source materials. An ongoing challenge when using BCT taxonomies relates to the uncertain intensity of each BCT[18]. Future research is needed to tackle this issue including guidance on how to code and report BCT intensity and dose received by participants.

## Conclusions

Our study provides important insight into the methods researchers have used when applying BCT taxonomies in childhood obesity prevention interventions. We found substantial variation in the methods used to apply BCT taxonomies and to report BCT-related methods and results. There was a paucity of detail reported in how BCTs were selected in studies applying BCT taxonomies prospectively. We propose a need for best practice reporting guidance to standardise processes of applying BCT taxonomies and improve the detail reported in publications. We discuss several of the ongoing challenges in how BCT taxonomies are applied and provide direction for future research in this important methods area.

## Data Availability

All data produced in the present study are available upon reasonable request to the authors

## DECLARATION OF INTEREST STATEMENT

### Author contributions

Conceptualization: BJJ and ALS. Methodology - Draft: DC and BAB. Methodology – Review & Editing: ALS, SY, KEH, RKH, ACW, BJJ. Investigation: DC, BAB, SY, BJ. Writing – Original Draft: BJJ, DC, BAB. Writing – Review & Editing: ALS, SY, KEH, RKH, ACW. Supervision: BJJ, ALS, KEH. All authors approved the final version of the manuscript.

## Footnotes

1 Behaviour change techniques

