## Supplementary files for "Exploring the application of behaviour change technique taxonomies in childhood obesity prevention interventions: A systematic scoping review"

**SUPPLEMENTARY ONLINE MATERIALS**

**Supplementary File 1:** Preferred Reporting Items for Systematic reviews and Meta-Analyses extension for Scoping Reviews (PRISMA-ScR) Checklist

| **SECTION** | **ITEM** | **PRISMA-ScR CHECKLIST ITEM** | **REPORTED ON PAGE #** |
| --- | --- | --- | --- |
| **TITLE** | | | |
| Title | 1 | Identify the report as a scoping review. | 1 |
| **ABSTRACT** | | | |
| Structured summary | 2 | Provide a structured summary that includes (as applicable): background, objectives, eligibility criteria, sources of evidence, charting methods, results, and conclusions that relate to the review questions and objectives. | 2 |
| **INTRODUCTION** | | | |
| Rationale | 3 | Describe the rationale for the review in the context of what is already known. Explain why the review questions/objectives lend themselves to a scoping review approach. | 3 |
| Objectives | 4 | Provide an explicit statement of the questions and objectives being addressed with reference to their key elements (e.g., population or participants, concepts, and context) or other relevant key elements used to conceptualize the review questions and/or objectives. | 3-4 |
| **METHODS** | | | |
| Protocol and registration | 5 | Indicate whether a review protocol exists; state if and where it can be accessed (e.g., a Web address); and if available, provide registration information, including the registration number. | 4 |
| Eligibility criteria | 6 | Specify characteristics of the sources of evidence used as eligibility criteria (e.g., years considered, language, and publication status), and provide a rationale. | 4 |
| Information sources* | 7 | Describe all information sources in the search (e.g., databases with dates of coverage and contact with authors to identify additional sources), as well as the date the most recent search was executed. | 4 |
| Search | 8 | Present the full electronic search strategy for at least 1 database, including any limits used, such that it could be repeated. | 4 |
| Selection of sources of evidence† | 9 | State the process for selecting sources of evidence (i.e., screening and eligibility) included in the scoping review. | 5 |
| Data charting process‡ | 10 | Describe the methods of charting data from the included sources of evidence (e.g., calibrated forms or forms that have been tested by the team before their use, and whether data charting was done independently or in duplicate) and any processes for obtaining and confirming data from investigators. | 5 |
| Data items | 11 | List and define all variables for which data were sought and any assumptions and simplifications made. | 5 |
| Critical appraisal of individual sources of evidence§ | 12 | If done, provide a rationale for conducting a critical appraisal of included sources of evidence; describe the methods used and how this information was used in any data synthesis (if appropriate). | N/A |
| Synthesis of results | 13 | Describe the methods of handling and summarizing the data that were charted. | 6 |
| **RESULTS** | | | |
| Selection of sources of evidence | 14 | Give numbers of sources of evidence screened, assessed for eligibility, and included in the review, with reasons for exclusions at each stage, ideally using a flow diagram. | 6-7, Fig 1 |
| Characteristics of sources of evidence | 15 | For each source of evidence, present characteristics for which data were charted and provide the citations. | 8-11, Suppl Files |
| Critical appraisal within sources of evidence | 16 | If done, present data on critical appraisal of included sources of evidence (see item 12). | N/A |
| Results of individual sources of evidence | 17 | For each included source of evidence, present the relevant data that were charted that relate to the review questions and objectives. | 8-11, Suppl Files |
| Synthesis of results | 18 | Summarize and/or present the charting results as they relate to the review questions and objectives. | 8-11, Table 2 and 3 |
| **DISCUSSION** | | | |
| Summary of evidence | 19 | Summarize the main results (including an overview of concepts, themes, and types of evidence available), link to the review questions and objectives, and consider the relevance to key groups. | 11-13 |
| Limitations | 20 | Discuss the limitations of the scoping review process. | 13 |
| Conclusions | 21 | Provide a general interpretation of the results with respect to the review questions and objectives, as well as potential implications and/or next steps. | 14 |
| **FUNDING** | | | |
| Funding | 22 | Describe sources of funding for the included sources of evidence, as well as sources of funding for the scoping review. Describe the role of the funders of the scoping review. | 14 |

JBI = Joanna Briggs Institute; PRISMA-ScR = Preferred Reporting Items for Systematic reviews and Meta-Analyses extension for Scoping Reviews.

*From:* Tricco AC, Lillie E, Zarin W, O'Brien KK, Colquhoun H, Levac D, et al. PRISMA Extension for Scoping Reviews (PRISMAScR): Checklist and Explanation. Ann Intern Med. 2018;169:467–473. [doi: 10.7326/M18-0850](http://annals.org/aim/fullarticle/2700389/prisma-extension-scoping-reviews-prisma-scr-checklist-explanation).

**Supplementary File 2:** Search strategy for all database searches

Ovid (Medline) 1946 to February 13, 2021

| Health condition: Overweight/obesity and related behaviours | 1 | Pediatric obesity/ or Obesity/ or Obesity, Abdominal/ or Obesity, Morbid/ |
| --- | --- | --- |
|  | 2 | Overweight/ |
|  | 3 | Weight gain/ |
|  | 4 | obes*.af |
|  | 5 | (overweight or over weight or over-weight).af |
|  | 6 | Exp Breastfeeding/ |
|  | 7 | Infant Nutritional Physiological Phenomena/ |
|  | 8 | Child Nutrition Sciences/ |
|  | 9 | Infant Food/ |
|  | 10 | ((child or toddler or infant$) adj1 (food or feeding or nutrition$)).tw. |
|  | 11 | ((responsive or complementary) adj1 feeding).ti,ab |
|  | 12 | ((diet* or nutrition) adj (modif* or strateg* or intervention* or advice or program* or class* or counsel* or educat* or instruct* or teach* or train* or guidance or lesson* or workshop* or module* or consultation* or session*)).ti,ab |
|  | 13 | (healthy eating).ti,ab |
|  | 14 | (fruit or vegetable*).ti,ab |
|  | 15 | (high fat* or low fat* or fatty food*).ti,ab |
|  | 16 | exp Exercise/ |
|  | 17 | exercis*.ti,ab |
|  | 18 | (physical activity or physical inactivity).ti,ab |
|  | 19 | Sedentary behavio?r.ti,ab |
|  | 20 | (screen time).ti,ab |
|  | 21 | Sleep/ |
|  | 22 | 1 or 2 or 3 or 4 or 5 or 6 or 7 or 8 or 9 or 10 or 11 or 12 or 13 or 14 or 15 or 16 or 17 or 18 or 19 or 20 or 21 |
| Phenomenon of interest: Behaviour change techniques | 23 | (BCT* or behaviour change strateg* or behaviour change technique* or behaviour* intervention* or behavior change strateg* or behavior change technique* or behavior* intervention*).af. |
| Population: children, adolescents and families | 24 | exp child/ or exp infant/ |
|  | 25 | (child* or infant* or pediatr* or paediatr* baby or toddler* or boys or girls or schoolchildren or school children).af. |
|  | 26 | (pregnan* or antenatal or parent or parent$1 or care giver or caregiver or guardian or family or families or mother$1 or father$1).af |
|  | 27 | 24 or 25 or 26 |
| Combine categories | 28 | 22 and 23 and 27 |
| Humans | 29 | (exp animals/ not humans.sh.) or (rat or rats or mouse or mice or rodent*).ti. |
|  | 30 | 28 not 29 |
| TOTAL | **1442** | |

EBM Reviews - Cochrane Central Register of Controlled Trials

| Health condition: Overweight/obesity and related behaviours | 1 | Pediatric obesity/ or Obesity/ or Obesity, Abdominal/ or Obesity, Morbid/ |
| --- | --- | --- |
|  | 2 | Overweight/ |
|  | 3 | Weight gain/ |
|  | 4 | obes*.af |
|  | 5 | (overweight or over weight or over-weight).af |
|  | 6 | exp Breastfeeding/ |
|  | 7 | Infant Nutritional Physiological Phenomena/ |
|  | 8 | Child Nutrition Sciences/ |
|  | 9 | Infant Food/ |
|  | 10 | ((child or toddler or infant$) adj1 (food or feeding or nutrition$)).tw. |
|  | 11 | ((responsive or complementary) adj1 feeding).ti,ab |
|  | 12 | ((diet* or nutrition) adj (modif* or strateg* or intervention* or advice or program* or class* or counsel* or educat* or instruct* or teach* or train* or guidance or lesson* or workshop* or module* or consultation* or session*)).ti,ab |
|  | 13 | (healthy eating).ti,ab |
|  | 14 | (fruit or vegetable*).ti,ab |
|  | 15 | (high fat* or low fat* or fatty food*).ti,ab |
|  | 16 | exp Exercise/ |
|  | 17 | exercis*.ti,ab |
|  | 18 | (physical activity or physical inactivity).ti,ab |
|  | 19 | sedentary behavio?r.ti,ab |
|  | 20 | (screen time).ti,ab |
|  | 21 | Sleep/ |
|  | 22 | 1 or 2 or 3 or 4 or 5 or 6 or 7 or 8 or 9 or 10 or 11 or 12 or 13 or 14 or 15 or 16 or 17 or 18 or 19 |
| Phenomenon of interest: Behaviour change techniques | 23 | (BCT* or behaviour change strateg* or behaviour change technique* or behaviour* intervention* or behavior change strateg* or behavior change technique* or behavior* intervention*).af. |
| Population: children, adolescents and families | 24 | exp child/ or exp infant/ |
|  | 25 | (child* or infant* or pediatr* or paediatr* baby or toddler* or boys or girls or schoolchildren or school children).af. |
|  | 26 | (pregnan* or antenatal or parent or parent$1 or care giver or caregiver or guardian or family or families or mother$1 or father$1).af |
|  | 27 | 24 or 25 or 26 |
| Combine categories | 28 | 22 and 23 and 27 |
| Humans | 29 | (exp animals/ not humans.sh.) or (rat or rats or mouse or mice or rodent*).ti. |
|  | 30 | 28 not 29 |
| TOTAL | **683** | |

EBM Reviews - Cochrane Database of Systematic Reviews 2005 to February 10, 2021

| Health condition: Overweight/obesity and related behaviours | 1 | Pediatric obesity/ or Obesity/ or Obesity, Abdominal/ or Obesity, Morbid/ |
| --- | --- | --- |
|  | 2 | Overweight/ |
|  | 3 | Weight gain/ |
|  | 4 | obes*.af |
|  | 5 | (overweight or over weight or over-weight).af |
|  | 6 | exp Breastfeeding/ |
|  | 7 | Infant Nutritional Physiological Phenomena/ |
|  | 8 | Child Nutrition Sciences/ |
|  | 9 | Infant Food/ |
|  | 10 | ((child or toddler or infant$) adj1 (food or feeding or nutrition$)).tw. |
|  | 11 | ((responsive or complementary) adj1 feeding).ti,ab |
|  | 12 | ((diet* or nutrition) adj (modif* or strateg* or intervention* or advice or program* or class* or counsel* or educat* or instruct* or teach* or train* or guidance or lesson* or workshop* or module* or consultation* or session*)).ti,ab |
|  | 13 | (healthy eating).ti,ab |
|  | 14 | (fruit or vegetable*).ti,ab |
|  | 15 | (high fat* or low fat* or fatty food*).ti,ab |
|  | 16 | exp Exercise/ |
|  | 17 | exercis*.ti,ab |
|  | 18 | (physical activity or physical inactivity).ti,ab |
|  | 19 | sedentary behavio?r.ti,ab |
|  | 20 | (screen time).ti,ab |
|  | 21 | Sleep/ |
|  | 22 | 1 or 2 or 3 or 4 or 5 or 6 or 7 or 8 or 9 or 10 or 11 or 12 or 13 or 14 or 15 or 16 or 17 or 18 or 19 |
| Phenomenon of interest: Behaviour change techniques | 23 | (BCT* or behaviour change strateg* or behaviour change technique* or behaviour* intervention* or behavior change strateg* or behavior change technique* or behavior* intervention*).af. |
| Population: children, adolescents and families | 24 | exp child/ or exp infant/ |
|  | 25 | (child* or infant* or pediatr* or paediatr* baby or toddler* or boys or girls or schoolchildren or school children).af. |
|  | 26 | (pregnan* or antenatal or parent or parent$1 or care giver or caregiver or guardian or family or families or mother$1 or father$1).af |
|  | 27 | 24 or 25 or 26 |
| Combine categories | 28 | 22 and 23 and 27 |
| Humans | 29 | (exp animals/ not humans.sh.) or (rat or rats or mouse or mice or rodent*).ti. |
|  | 30 | 28 not 29 |
| TOTAL | **283** | |

Cochrane Library

| Health condition: Overweight/obesity and related behaviours | 1 | Pediatric obesity OR Obesity OR Obesity, Abdominal OR Obesity, Morbid |
| --- | --- | --- |
|  | 2 | Overweight |
|  | 3 | Weight gain |
|  | 4 | obes* |
|  | 5 | overweight OR over weight OR over-weight |
|  | 6 | MeSH descriptor: [Breast Feeding] explode all trees |
|  | 7 | Infant Nutritional Physiological Phenomena |
|  | 8 | Child Nutrition Sciences |
|  | 9 | Infant Food |
|  | 10 | (child OR toddler OR infant$) adj1 (food OR feeding OR nutrition$) |
|  | 11 | (responsive OR complementary) adj1 feeding |
|  | 12 | (diet* OR nutrition) adj (modif* OR strateg* OR intervention* OR advice or program* OR class* OR counsel* OR educat* OR instruct* OR teach* OR train* OR guidance OR lesson* OR workshop* OR module* OR consultation* OR session*) |
|  | 13 | healthy eating |
|  | 14 | fruit OR vegetable* |
|  | 15 | high fat* OR low fat* OR fatty food* |
|  | 16 | exercis* |
|  | 17 | MeSH descriptor: [Exercise] explode all trees |
|  | 18 | physical activity OR physical inactivity |
|  | 19 | sedentary behavior |
|  | 20 | screen time |
|  | 21 | Sleep |
|  | 22 | {OR #1-#21} |
| Phenomenon of interest: Behaviour change techniques | 23 | (BCT* OR behaviour change strateg* OR behaviour change technique* OR behaviour* intervention* OR behavior change strateg* OR behavior change technique* OR behavior* intervention*) |
| Population: children, adolescents and families | 24 | exp child/ OR exp infant/ |
|  | 25 | (child* or infant* or pediatr* or paediatr* baby or toddler* or boys or girls or schoolchildren or school children).af. |
|  | 26 | (pregnan* OR antenatal OR parent OR parent$1 OR care giver OR caregiver OR guardian OR family OR families OR mother$1 OR father$1) |
|  | 27 | {OR #24-#26} |
| Combine categories | 28 | #22 AND #23 AND #27 |
| Humans | 29 | exp animals NOT humans |
|  | 30 | rat or rats or mouse or mice or rodent* |
|  | 31 | {OR #29-#30} |
|  | 32 | #28 NOT #31 |
| TOTAL | **120** | |

PsychInfo

| Health condition: Overweight/obesity and related behaviours | 1 | Pediatric obesity/ or Obesity/ or Obesity, Abdominal/ or Obesity, Morbid/ |
| --- | --- | --- |
|  | 2 | Overweight/ |
|  | 3 | Weight gain/ |
|  | 4 | obes*.af |
|  | 5 | (overweight or over weight or over-weight).af |
|  | 6 | exp Breastfeeding/ |
|  | 7 | Infant Nutritional Physiological Phenomena/ |
|  | 8 | Child Nutrition Sciences/ |
|  | 9 | Infant Food/ |
|  | 10 | ((child or toddler or infant$) adj1 (food or feeding or nutrition$)).tw. |
|  | 11 | ((responsive or complementary) adj1 feeding).ti,ab |
|  | 12 | ((diet* or nutrition) adj (modif* or strateg* or intervention* or advice or program* or class* or counsel* or educat* or instruct* or teach* or train* or guidance or lesson* or workshop* or module* or consultation* or session*)).ti,ab |
|  | 13 | (healthy eating).ti,ab |
|  | 14 | (fruit or vegetable*).ti,ab |
|  | 15 | (high fat* or low fat* or fatty food*).ti,ab |
|  | 16 | exp Exercise/ |
|  | 17 | exercis*.ti,ab |
|  | 18 | (physical activity or physical inactivity).ti,ab |
|  | 19 | sedentary behavio?r.ti,ab |
|  | 20 | (screen time).ti,ab |
|  | 21 | Sleep/ |
|  | 22 | 1 or 2 or 3 or 4 or 5 or 6 or 7 or 8 or 9 or 10 or 11 or 12 or 13 or 14 or 15 or 16 or 17 or 18 or 19 or 20 or 21 |
| Phenomenon of interest: Behaviour change techniques | 23 | (BCT* or behaviour change strateg* or behaviour change technique* or behavior change strateg* or behavior change technique*).af. |
| Population: children, adolescents and families | 24 | exp child/ or exp infant/ |
|  | 25 | (child* or infant* or pediatr* or paediatr* baby or toddler* or boys or girls or schoolchildren or school children).af. |
|  | 26 | (pregnan* or antenatal or parent or parent$1 or care giver or caregiver or guardian or family or families or mother$1 or father$1).af |
|  | 27 | 24 or 25 or 26 |
| Combine categories | 28 | 22 and 23 and 27 |
| Humans | 29 | (exp animals/ not humans.sh.) or (rat or rats or mouse or mice or rodent*).ti. |
|  | 30 | 28 not 29 |

Embase

| Health condition: Overweight/obesity and related behaviours | 1 | Pediatric obesity/ or Obesity/ or Obesity, Abdominal/ or Obesity, Morbid/ |
| --- | --- | --- |
|  | 2 | Overweight/ |
|  | 3 | Weight gain/ |
|  | 4 | obes*.af |
|  | 5 | (overweight or over weight or over-weight).af |
|  | 6 | exp Breastfeeding/ |
|  | 7 | Infant Nutritional Physiological Phenomena/ |
|  | 8 | Child Nutrition Sciences/ |
|  | 9 | Infant Food/ |
|  | 10 | ((child or toddler or infant$) adj1 (food or feeding or nutrition$)).tw. |
|  | 11 | ((responsive or complementary) adj1 feeding).ti,ab |
|  | 12 | ((diet* or nutrition) adj (modif* or strateg* or intervention* or advice or program* or class* or counsel* or educat* or instruct* or teach* or train* or guidance or lesson* or workshop* or module* or consultation* or session*)).ti,ab |
|  | 13 | (healthy eating).ti,ab |
|  | 14 | (fruit or vegetable*).ti,ab |
|  | 15 | (high fat* or low fat* or fatty food*).ti,ab |
|  | 16 | exp Exercise/ |
|  | 17 | exercis*.ti,ab |
|  | 18 | (physical activity or physical inactivity).ti,ab |
|  | 19 | sedentary behavio?r.ti,ab |
|  | 20 | (screen time).ti,ab |
|  | 21 | Sleep/ |
|  | 22 | 1 or 2 or 3 or 4 or 5 or 6 or 7 or 8 or 9 or 10 or 11 or 12 or 13 or 14 or 15 or 16 or 17 or 18 or 19 or 20 or 21 |
| Phenomenon of interest: Behaviour change techniques | 23 | (BCT* or behaviour change strateg* or behaviour change technique* or behaviour* intervention* or behavior change strateg* or behavior change technique* or behavior* intervention*).af. |
| Population: children, adolescents and families | 24 | exp child/ or exp infant/ |
|  | 25 | (child* or infant* or pediatr* or paediatr* baby or toddler* or boys or girls or schoolchildren or school children).af. |
|  | 26 | (pregnan* or antenatal or parent or parent$1 or care giver or caregiver or guardian or family or families or mother$1 or father$1).af |
|  | 27 | 24 or 25 or 26 |
| Combine categories | 28 | 22 and 23 and 27 |
| Humans | 29 | (exp animals/ not humans.sh.) or (rat or rats or mouse or mice or rodent*).ti. |
|  | 30 | 28 not 29 |

CINHAL

| Health condition: Overweight/obesity and related behaviours | 1 | (MH "Obesity, Morbid") OR (MH "Pediatric Obesity") OR (MH "Obesity") |
| --- | --- | --- |
|  | 2 | (MH "Weight Gain") |
|  | 3 | TX obes* |
|  | 4 | TX overweight OR TX "over weight" OR TX over-weight |
|  | 5 | (MH "Breast Feeding+") |
|  | 6 | (MH "Infant Nutritional Physiology+") |
|  | 7 | (MH "Child Nutritional Physiology+") |
|  | 8 | (MH "Infant Food+") |
|  | 9 | TI ( ((child or toddler or infant$) N2 (food or feeding or nutrition$)) ) OR AB ( ((child or toddler or infant$) N2 (food or feeding or nutrition$)) ) |
|  | 10 | TI ( ((responsive or complementary) N2 feeding) ) OR AB ( ((responsive or complementary) N2 feeding) ) |
|  | 11 | TI ( ((diet* or nutrition) N2 (modif* or strateg* or intervention* or advice or program* or class* or counsel* or educat* or instruct* or teach* or train* or guidance or lesson* or workshop* or module* or consultation* or session*)) ) AND AB ( ((diet* or nutrition) N2 (modif* or strateg* or intervention* or advice or program* or class* or counsel* or educat* or instruct* or teach* or train* or guidance or lesson* or workshop* or module* or consultation* or session*)) ) |
|  | 12 | TI "healthy eating" OR AB "healthy eating" |
|  | 13 | TI ( fruit or vegetable* ) OR AB ( fruit or vegetable* ) |
|  | 14 | TI ( high fat* or low fat* or fatty food* ) OR AB ( high fat* or low fat* or fatty food* ) |
|  | 15 | (MH "Exercise+") |
|  | 16 | TI exercis* OR AB exercis* |
|  | 17 | TI ( (physical activity or physical inactivity) ) OR AB ( (physical activity or physical inactivity) ) |
|  | 18 | TI "sedentary behavio?r" OR AB "sedentary behavio?r" |
|  | 19 | TI "screen time" OR AB "screen time" |
|  | 20 | (MH "Sleep+") |
|  | 21 | S1 or S2 or S3 or S4 or S5 or S6 or S7 or S8 or S9 or S10 or S11 or S12 or S13 or S14 or S15 or S16 or S17 or S18 or S19 or S20 |
|  | 22 | TX (BCT* or "behaviour* change strateg*" or "behaviour* change technique*" or "behaviour* intervention"* or "behavior* change strateg*" or "behavior* change technique*" or "behavior* intervention*") |
| Phenomenon of interest: Behaviour change techniques | 23 | (MH "Child+") |
| Population: children, adolescents and families | 24 | (MH "Infant+") |
|  | 25 | TX (child* or infant* or pediatr* or paediatr* or baby or toddler* or boys or girls or schoolchildren or "school children") |
|  | 26 | TX (pregnan* or antenatal or parent or parent$1 or care giver or caregiver or guardian or family or families or mother$1 or father$1) |
|  | 27 | S23 or S24 or S25 or S26 |
| Combine categories | 28 | S21 and S22 and S27 |
| Humans | 29 | (MH "Animals+") |
|  | 30 | S28 NOT S29 |

PROSPERO, until March 7, 2021

| Line |  |
| --- | --- |
| #1 | obesity |
| #2 | abdominal obesity |
| #3 | morbid obesity |
| #4 | overweight |
| #5 | over weight |
| #6 | over-weight |
| #7 | weight gain |
| #8 | obes* |
| #9 | breastfeeding |
| #10 | Infant Nutritional Physiological Phenomena |
| #11 | Child Nutrition Sciences |
| #12 | Infant Food |
| #13 | ((child or toddler or infant$) adj1 (food or feeding or nutrition$)) |
| #14 | ((responsive or complementary) adj1 feeding) |
| #15 | healthy eating |
| #16 | fruit OR vegetable* |
| #17 | (high fat* or low fat* or fatty food*) |
| #18 | exercis* |
| #19 | (physical activity or physical inactivity) |
| #20 | sedentary behaviour |
| #21 | screen time |
| #22 | sleep |
| #23 | #1 OR #2 OR #3 OR #4 OR #5 OR #6 OR #7 OR #8 OR #9 OR #10 OR #11 OR #12 OR #13 OR #14 OR #15 OR #16 OR #17 OR #18 OR #19 OR #20 OR #21 OR #22 |
| #24 | behaviour change technique* |
| #25 | BCT |
| #26 | behaviour change strateg* |
| #27 | #24 OR #25 OR #26 |
| #28 | (child* or infant* or?pediatr* or?paediatr* baby or toddler* or boys or girls or schoolchildren or school children) |
| #29 | (pregnan* or antenatal or parent or parent$1 or care giver or caregiver or guardian or family or families or mother$1 or father$1) |
| #30 | #28 OR #29 |
| #31 | #23 AND #27 AND #30 |

**Supplementary File 3:** References for studies included in the review

**Prospective studies**

Arredondo et al (2018)

Arredondo EM, Ayala GX, Soto S, Slymen DJ, Horton LA, Parada H, et al. Latina mothers as agents of change in children’s eating habits: findings from the randomized controlled trial Entre Familia: Reflejos de Salud. International Journal of Behavioral Nutrition and Physical Activity. 2018;15(1):95.

Burton et al (2021)

Burton W, Sahota P, Twiddy M, Brown J, Bryant M. The Development of a Multilevel Intervention to Optimise Participant Engagement with an Obesity Prevention Programme Delivered in UK children's Centres. Prev Sci. 2021;22(3):345-56.

Clarke et al (2020) and Jolley et al (2018)

Clarke JL, Ingram J, Johnson D, Thomson G, Trickey H, Dombrowski SU, et al. The ABA intervention for improving breastfeeding initiation and continuation: Feasibility study results. Matern Child Nutr. 2020;16(1):e12907.

Jolly K, Ingram J, Clarke J, Johnson D, Trickey H, Thomson G, et al. Protocol for a feasibility trial for improving breast feeding initiation and continuation: assets-based infant feeding help before and after birth (ABA). BMJ Open. 2018;8(1):e019142.

Downing et al (2018)

Downing KL, Salmon J, Hinkley T, Hnatiuk JA, Hesketh KD. Feasibility and Efficacy of a Parent-Focused, Text Message-Delivered Intervention to Reduce Sedentary Behavior in 2- to 4-Year-Old Children (Mini Movers): Pilot Randomized Controlled Trial. JMIR Mhealth Uhealth. 2018;6(2):e39.

Draper et al (2019)

Draper CE, Howard SJ, Rochat TJ. Feasibility and acceptability of a home-based intervention to promote nurturing interactions and healthy behaviours in early childhood: The Amagugu Asakhula pilot study. Child Care Health Dev. 2019;45(6):823-31.

Duncan et al (2011)

Duncan S, McPhee J, Schluter P, Zinn C, Smith R, Schofield G. Efficacy of a compulsory homework programme for increasing physical activity and healthy eating in children: the healthy homework pilot study. International Journal of Behavioral Nutrition and Physical Activity. 2011;8(127).

Duncan et al (2019)

Duncan S, Stewart T, McPhee J, Borotkanics R, Prendergast K, Zinn C, et al. Efficacy of a compulsory homework programme for increasing physical activity and improving nutrition in children: a cluster randomised controlled trial. International Journal of Behavioral Nutrition and Physical Activity. 2019;16(1):80.

Espinosa-Curiel et al (2020)

Espinosa-Curiel IE, Pozas-Bogarin EE, Lozano-Salas JL, Martinez-Miranda J, Delgado-Perez EE, Estrada-Zamarron LS. Nutritional Education and Promotion of Healthy Eating Behaviors Among Mexican Children Through Video Games: Design and Pilot Test of FoodRateMaster. JMIR Serious Games. 2020;8(2):e16431.

Fisher et al (2019)

Fisher JO, Serrano EL, Foster GD, Hart CN, Davey A, Bruton YP, et al. Efficacy of a food parenting intervention for mothers with low income to reduce preschooler’s solid fat and added sugar intakes: a randomized controlled trial. International Journal of Behavioral Nutrition and Physical Activity. 2019;16(1):6.

Fletcher et al (2013) and Wyse et al (2014)

Fletcher A, Wolfenden L, Wyse R, Bowman J, McElduff P, Duncan S. A randomised controlled trial and mediation analysis of the ‘Healthy Habits’, telephone-based dietary intervention for preschool children. International Journal of Behavioral Nutrition and Physical Activity. 2013;10(1):43.

Wyse R, Campbell KJ, Brennan L, Wolfenden L. A cluster randomised controlled trial of a telephone-based intervention targeting the home food environment of preschoolers (The Healthy HabitsTrial): the effect on parent fruit and vegetable consumption. International Journal of Behavioral Nutrition and Physical Activity. 2014;11(1):144.

Fulkerson et al (2015)

Fulkerson JA, Friend S, Flattum C, Horning M, Draxten M, Neumark-Sztainer D, et al. Promoting healthful family meals to prevent obesity: HOME Plus, a randomized controlled trial. International Journal of Behavioral Nutrition and Physical Activity. 2015;12(1):154.

Gholami et al (2015)

Gholami M, Wiedemann A, Knoll N, Schwarzer R. Mothers improve their daughters’ vegetable intake: A randomized controlled trial. Psychology, Health & Medicine. 2015;20(1):1-7.

Hammersley et al (2020) (protocol)

Hammersley ML, Wyse RJ, Jones RA, Wolfenden L, Yoong S, Stacey F, et al. Translation of two healthy eating and active living support programs for parents of 2–6 year old children: a parallel partially randomised preference trial protocol (the ‘time for healthy habits’ trial). BMC Public Health. 2020;20(1):636.

Lakshman et al (2014, 2018)

Lakshman R, Griffin S, Hardeman W, Schiff A, Kinmonth AL, Ong KK. Using the Medical Research Council framework for the development and evaluation of complex interventions in a theory-based infant feeding intervention to prevent childhood obesity: the baby milk intervention and trial. J Obes. 2014;2014:646504.

Lakshman R, Sharp SJ, Whittle F, Schiff A, Hardeman W, Irvine L, et al. Randomised controlled trial of a theory-based behavioural intervention to reduce formula milk intake. Arch Dis Child. 2018;103(11):1054-60.

Lin et al (2016)

Lin M, Pan L-p, Han J, Li L, Jiang J-x, Jin R-m. Behavioral intervention reduces unhealthy eating behaviors in preschool children via a behavior card approach. Journal of Huazhong University of Science and Technology [Medical Sciences]. 2016;36(6):895-903.

Lloyd et al (2011)

Lloyd J, Logan S, Greaves C, Wyatt K. Evidence, theory and context - using intervention mapping to develop a school-based intervention to prevent obesity in children. international Journal of Behavioral Nutrition and Physical Activity. 2011;8(73).

Majumdar et al (2013)

Majumdar D, Koch PA, Lee H, Contento IR, Islas-Ramos AD, Fu D. "Creature-101": A Serious Game to Promote Energy Balance-Related Behaviors Among Middle School Adolescents. Games Health J. 2013;2(5):280-90.

Marshall et al (2021)

Marshall S, Taki S, Love P, Laird Y, Kearney M, Tam N, et al. The process of culturally adapting the Healthy Beginnings early obesity prevention program for Arabic and Chinese mothers in Australia. BMC Public Health. 2021;21(1):284.

Martin et al (2015)

Martin R, Murtagh EM. Preliminary findings of Active Classrooms: An intervention to increase physical activity levels of primary school children during class time. Teaching and Teacher Education. 2015;52:113-27.

McSweeney et al (2017)

McSweeney L, Araújo-Soares V, Rapley T, Adamson A. A feasibility study with process evaluation of a preschool intervention to improve child and family lifestyle behaviours. BMC Public Health. 2017;17(1):248.

Morgan et al (2019)

Morgan PJ, Young MD, Barnes AT, Eather N, Pollock ER, Lubans DR. Engaging Fathers to Increase Physical Activity in Girls: The "Dads And Daughters Exercising and Empowered" (DADEE) Randomized Controlled Trial. Ann Behav Med. 2019;53(1):39-52.

Morrison et al (2013) and Yam et al (2012)

Morrison R, Reilly JJ, Penpraze V, Westgarth C, Ward DS, Mutrie N, et al. Children, parents and pets exercising together (CPET): exploratory randomised controlled trial. BMC Public Health 2013;13:1096

Yam PS, Morrison R, Penpraze V, Westgarth C, Ward DS, Mutrie N, et al. Children, parents, and pets exercising together (CPET) randomised controlled trial: study rationale, design, and methods. BMC Public Health. 2012;12:208.

Phillips et al (2018)

Phillips R, Copeland L, Grant A, Sanders J, Gobat N, Tedstone S, et al. Development of a novel motivational interviewing (MI) informed peer-support intervention to support mothers to breastfeed for longer. BMC Pregnancy Childbirth. 2018;18(1):90.

Po'e et al (2013)

Po'e EK, Heerman WJ, Mistry RS, Barkin SL. Growing Right Onto Wellness (GROW): a family-centered, community-based obesity prevention randomized controlled trial for preschool child-parent pairs. Contemp Clin Trials. 2013;36(2):436-49.

Powell et al (2016)

Powell E, Woodfield LA, Nevill AM. Increasing physical activity levels in primary school physical education: The SHARP Principles Model. Preventive Medicine Reports. 2016;3:7-13.

Roberts-Gray et al (2016)

Roberts-Gray C, Briley ME, Ranjit N, Byrd-Williams CE, Sweitzer SJ, Sharma SV, et al. Efficacy of the Lunch is in the Bag intervention to increase parents’ packing of healthy bag lunches for young children: a cluster-randomized trial in early care and education centers. International Journal of Behavioral Nutrition and Physical Activity. 2016;13(1):3.

Russell et al (2018)

Russell C, Denney-Wilson E, Laws R, Abbott G, Zheng M, Lymer S, et al. Impact of the Growing Healthy mHealth Program on Maternal Feeding Practices, Infant Food Preferences, and Satiety Responsiveness: Quasi-Experimental Study. JMIR Mhealth Uhealth. 2018;6(4):e77.

Skouteris et al (2016)

Skouteris H, Hill B, McCabe M, Swinburn B, Busija L. A parent-based intervention to promote healthy eating and active behaviours in pre-school children: evaluation of the MEND 2-4 randomized controlled trial. Pediatr Obes. 2016;11(1):4-10.

Smith et al (2018)

Smith, J.D., et al., The Family Check-Up 4 Health (FCU4Health): Applying Implementation Science Frameworks to the Process of Adapting an Evidence-Based Parenting Program for Prevention of Pediatric Obesity and Excess Weight Gain in Primary Care. Frontiers in Public Health, 2018. 6(293).

Taylor et al (2013)

Taylor NJ, Sahota P, Sargent J, Barber S, Loach J, Louch G, et al. Using intervention mapping to develop a culturally appropriate intervention to prevent childhood obesity: the HAPPY (Healthy and Active Parenting Programme for Early Years) study. International Journal of Behavioral Nutrition and Physical Activity. 2013;10(1):142.

Toomey et al (2020)

Toomey E, Matvienko-Sikar K, Doherty E, Harrington J, Hayes CB, Heary C, et al. A collaborative approach to developing sustainable behaviour change interventions for childhood obesity prevention: Development of the Choosing Healthy Eating for Infant Health (CHErIsH) intervention and implementation strategy. British Journal of Health Psychology. 2020;25(2):275-304.

Vaughn et al (2019) and Hennink-Kaminski et al (2018) (protocol)

Vaughn AE, Bartlett R, Luecking CT, Hennink-Kaminski H, Ward DS. Using a social marketing approach to develop Healthy Me, Healthy We: a nutrition and physical activity intervention in early care and education. Translational Behavioral Medicine. 2018;9(4):669-81.

Hennink-Kaminski H, Vaughn AE, Hales D, Moore RH, Luecking CT, Ward DS. Parent and child care provider partnerships: Protocol for the Healthy Me, Healthy We (HMHW) cluster randomized control trial. Contemporary Clinical Trials. 2018;64:49-57.

**Retrospective studies**

Anselma et al (2020) / Anselma et al (2016) (registration)

Anselma M, Chinapaw MJM, Kornet-van der Aa DA, Altenburg TM. Effectiveness and promising behavior change techniques of interventions targeting energy balance related behaviors in children from lower socioeconomic environments: A systematic review. PLoS One. 2020;15(9):e0237969.

Manou Anselma, Danielle van der Aa, Teatske Altenburg, Mai Chinapaw. A systematic review on the effectiveness of interventions aiming to improve physical activity, sedentary behavior or dietary behavior of children from lower socioeconomic environments. PROSPERO 2016 CRD42016052599

Azevedo et al (2019)

Azevedo LB, van Sluijs EMF, Moore HJ, Hesketh K. Determinants of change in accelerometer-assessed sedentary behaviour in children 0 to 6 years of age: A systematic review. Obes Rev. 2019;20(10):1441-64.

Busch et al (2017)

Busch V, Altenburg TM, Harmsen IA, Chinapaw MJ. Interventions that stimulate healthy sleep in school-aged children: a systematic literature review. Eur J Public Health. 2017;27(1):53-65.

Farre et al (2021) (registration)

Albert Farre, Louise Wallace, Anna Gavine, Joyce Marshall, Phyllis Buchanan, Angie Wade, Fiona Lynn, Jonathan West, Alison McFadden. Implementation of UK-relevant effective interventions to support women to breastfeed: a systematic review and mixed methods synthesis with embedded stakeholder engagement. PROSPERO 2021 CRD42021229769

Hendrie et al (2017)

Hendrie GA, Lease HJ, Bowen J, Baird DL, Cox DN. Strategies to increase children's vegetable intake in home and community settings: a systematic review of literature. Matern Child Nutr. 2017;13(1).

Hennessy et al (2019)

Hennessy M, Heary C, Laws R, van Rhoon L, Toomey E, Wolstenholme H, et al. The effectiveness of health professional-delivered interventions during the first 1000 days to prevent overweight/obesity in children: A systematic review. Obes Rev. 2019;20(12):1691-707.

JaKa et al (2019)

JaKa MM, French SA, Wolfson J, Jeffery RW, Lorencatto F, Michie S, et al. Understanding Outcomes in Behavior Change Interventions to Prevent Pediatric Obesity: The Role of Dose and Behavior Change Techniques. Health Educ Behav. 2019;46(2):312-21.

Jaka et al (2021)

JaKa MM, Wood C, Veblen-Mortenson S, Moore SM, Matheson D, Stevens J, et al. Applying the Behavior Change Technique Taxonomy to Four Multicomponent Childhood Obesity Interventions. West J Nurs Res. 2020:193945920954782.

Johnson et al (2018)

Johnson BJ, Zarnowiecki D, Hendrie GA, Mauch CE, Golley RK. How to reduce parental provision of unhealthy foods to 3- to 8-year-old children in the home environment? A systematic review utilizing the Behaviour Change Wheel framework. Obes Rev. 2018;19(10):1359-70.

Johnson et al (2020) (protocol pre-print), Seidler et al (2020) (registration)

Johnson BJ, Hunter KE, Golley RK, Chadwick P, Barba A, Aberoumand M, et al. Unpacking the behavioural components and delivery features of early childhood obesity prevention interventions in the TOPCHILD Collaboration: a systematic review and intervention coding protocol. medRxiv. 2020:2020.12.17.20248435.

Anna Lene Seidler, Kylie Hunter, Brittany Johnson, Angie Barba, Sol Libesman. Transforming Obesity Prevention in CHILDren (TOPCHILD): a systematic review and individual participant data meta-analysis to evaluate behavioural interventions for the prevention of very early childhood obesity. PROSPERO 2020 CRD42020177408

Kassianos et al (2019)

Kassianos AP, Ward E, Rojas-Garcia A, Kurti A, Mitchell FC, Nostikasari D, et al. A systematic review and meta-analysis of interventions incorporating behaviour change techniques to promote breastfeeding among postpartum women. Health Psychol Rev. 2019;13(3):344-72.

Keys et al (2020a) (registration)

Elizabeth Keys, Christine Cassidy, Shelly Weiss, Penny Corkum, Michelle Johnson, Leeanne Richardson, Marsha` Campbell-Yeo. Effectiveness of eHealth programs for parents of infants aged 1 year and under: A systematic review and best-fit framework synthesis of behavior change components.. PROSPERO 2020 CRD42020165339

Keys et al (2020b) (registration)

Elizabeth Keys, Christine Cassidy, Matt Orr, Ruth Shelton, Kaitlyn Butterfield, Fallon Cook, Shelly Weiss, Penny Corkum. Behavior change components of interventions with a behavioral sleep outcome for infants aged 1 year and under: A systematic review and meta-analysis using the Behavior Change Wheel.. PROSPERO 2020 CRD42020153160

Lewis et al 2019 (registration) and Lewis et al (2021)

Lewis L, Povey R, Rose S, Cowap L, Semper H, Carey A, et al. What behavior change techniques are associated with effective interventions to reduce screen time in 0-5 year olds? A narrative systematic review. Prev Med Rep. 2021;23:101429.

Lesley Lewis, Rachel Povey, Sarah Rose, David Clark-Carter, Heather Semper. What behaviour change techniques are associated with effective interventions to reduce screen time in 0-5 year olds? A systematic review. PROSPERO 2019 CRD42019129235

Masteller et al (2017)

Masteller B, Sirard J, Freedson P. The Physical Activity Tracker Testing in Youth (P.A.T.T.Y.) Study: Content Analysis and Children’s Perceptions. JMIR Mhealth Uhealth. 2017;5(4).

Matvienko-Sikar et al (2019) and Matvienko-Sikar et al (2016) (registration)

Matvienko-Sikar K, Toomey E, Delaney L, Flannery C, McHugh S, McSharry J, et al. Behaviour change techniques and theory use in healthcare professional-delivered infant feeding interventions to prevent childhood obesity: a systematic review. Health Psychol Rev. 2019;13(3):277-94.

Karen Matvienko-Sikar, Elaine Toomey, Lisa Delaney, Janas Harrington, Molly Byrne, Patricia Kearney. Systematic review of interventions influencing early infant feeding practices. PROSPERO 2016 CRD42016033492

Mauch et al (2018)

Mauch CE, Wycherley TP, Laws RA, Johnson BJ, Bell LK, Golley RK. Mobile Apps to Support Healthy Family Food Provision: Systematic Assessment of Popular, Commercially Available Apps. JMIR Mhealth Uhealth. 2018;6(12):e11867.

Seidler et al (2020)

Seidler AL, Hunter KE, Johnson BJ, Ekambareshwar M, Taki S, Mauch CE, et al. Understanding, comparing and learning from the four EPOCH early childhood obesity prevention interventions: A multi-methods study. Pediatr Obes. 2020;15(11):e12679.

Smith et al (2018) (as above)

Smith, J.D., et al., The Family Check-Up 4 Health (FCU4Health): Applying Implementation Science Frameworks to the Process of Adapting an Evidence-Based Parenting Program for Prevention of Pediatric Obesity and Excess Weight Gain in Primary Care. Frontiers in Public Health, 2018. 6(293).

vander Kruk et al (2013)

van der Kruk JJ, Kortekaas F, Lucas C, Jager-Wittenaar H. Obesity: a systematic review on parental involvement in long-term European childhood weight control interventions with a nutritional focus. Obes Rev. 2013;14(9):745-60.

Vargas-Garcia et al (2015) (protocol) and Vargas-Garcia et al (2017)

Vargas-Garcia EJ, El Evans C, Cade JE. Impact of interventions to reduce sugar-sweetened beverage intake in children and adults: a protocol for a systematic review and meta-analysis. Syst Rev. 2015;4:17.

Vargas-Garcia EJ, Evans CEL, Prestwich A, Sykes-Muskett BJ, Hooson J, Cade JE. Interventions to reduce consumption of sugar-sweetened beverages or increase water intake: evidence from a systematic review and meta-analysis. Obes Rev. 2017;18(11):1350-63.

Webb Girard et al (2020)

Webb Girard A, Waugh E, Sawyer S, Golding L, Ramakrishnan U. A scoping review of social-behaviour change techniques applied in complementary feeding interventions. Matern Child Nutr. 2020;16(1):e12882.

Zhang et al (2020) (registration)

Chun-Qing Zhang, Ru Zhang, Pak-Kwong Chung, Sam Liu, Ryan Rhodes. Family-based interventions to increase physical activity in children 3-12 years old: a systematic review, meta-analysis, and realist synthesis. PROSPERO 2020 CRD42020173324

**Supplementary Table 1:** Researcher methods used to code BCT in childhood obesity prevention prospective studies (n=32)

| **Author and Year**  **Location**  **Study design** | **Population** | **Target behaviour(s)** | **Reported BCTs** | **Selection of BCTs process** |
| --- | --- | --- | --- | --- |
| Arredondo et al (2018)  USA  RCT | N=361  Mothers with children aged 7-13y (Mexican living in California)    **Target Population of BCTs:** Mothers and children | Dietary intake | **Total:** Not reported    **Taxonomy:** Not reported    **BCTs reported with number and label:** N | **BCT Training:** Not reported  **Selection of BCTs:**  Social Cognitive Theory guided selection of BCTS |
| Burton et al (2021)  UK  Intervention development | N/A  Caregivers of young children    **Target Population of BCTs:** Commissioner, managers, childcare staff, parents who have attended intervention, intervention facilitators | Increasing engagement in an obesity prevention intervention:    Dietary intake  Physical activity | **Total:** 7    **Taxonomy:** BCTTv1    **BCTs reported with number and label:** Y | **BCT Training:** Not reported  **Selection of BCTs:**  Behaviour change wheel three stage process by the intervention development team:   - Key findings from a literature review and ethnography study presented to the team, developed target behaviours and completed a COM-B behavioural analysis - APEASE criteria to select intervention functions - APEASE criteria to select BCTs     Intervention development team included experts in intervention development, obesity, applied health research and behaviour change, local government representative; a HENRY parent; and the chief executive of HENRY. Parent advisory group consulted. |
| Clarke et al (2020) and Jolley et al (2018)  UK  Pilot RCT | N=103  Women pregnant with their first child    **Target Population of BCTs:** Pregnant women | Infant feeding | **Total:** 15    **Taxonomy:** BCTTv1    **BCTs reported with number and label:** Y | **BCT Training:** Not reported  **Selection of BCTs:**  Behaviour change wheel, including COM-B behavioural analysis, Theoretical Domains Framework to select intervention functions and BCTs. APEASE criteria used to select BCTs.    Information from systematic reviews, surveys, qualitative studies and discussions with end users to design the intervention. |
| Downing et al (2018)  Australia  Pilot RCT | N=57  Parents with children aged 2-4y    **Target Population of BCTs:** Parents | Sedentary behaviour | **Total:** 4    **Taxonomy:** CALO-RE    **BCTs reported with number and label:** N | **BCT Training:** Not reported  **Selection of BCTs:**  Intervention development was guided by the CALO-RE taxonomy and social cognitive theory. |
| Draper et al (2019)  South Africa  Pilot study (no control) | N=20  Caregivers of preschool children    **Target Population of BCTs:** Caregivers (mother, aunt, other) | Dietary intake  Physical activity  Sedentary behaviour  Sleep | **Total:** 14    **Taxonomy:** BCTTv1    **BCTs reported with number and label:** N | **BCT Training:** Not reported  **Selection of BCTs:** Not reported |
| Duncan et al (2011)  New Zealand  Pilot RCT | N=97  Children aged 9-11y    **Target Population of BCTs:** Children | Dietary intake  Physical activity  Sedentary behaviour | **Total:** 13    **Taxonomy:** Abraham et al 2008    **BCTs reported with number and label:** N | **BCT Training:** Not reported  **Selection of BCTs:** Not reported |
| Duncan et al (2019)  New Zealand  Cluster RCT | N=675  Children aged 7-10y    **Target Population of BCTs:** Children | Dietary intake  Physical activity | **Total:** Not reported    **Taxonomy:** Abraham et al 2008    **BCTs reported with number and label:** N | **BCT Training:** Not reported  **Selection of BCTs:**  Intervention development by advisory group including experts in health and education professionals. Regular input from children and parents. |
| Espinosa-Curiel et al (2020)  Mexico  Non-randomised trial | N=60  Children aged 8-10y    **Target Population of BCTs:** Children | Dietary intake  Physical activity | **Total:** 23    **Taxonomy:** BCTTv1    **BCTs reported with number and label:** N | **BCT Training:** Not reported  **Selection of BCTs:**  Video game intervention was developed using a two-step process:  Step 1:   - Literature review - Design sessions with two nutritionists and a psychologist to establish or adjust learning objectives, target behaviours, behaviour change objectives and BCTs   Step 2:   - Design sessions with two nutritionists, one psychologist, one expert on human-computer interaction, and two video game designers, to develop game including to implement the selected BCTs into the gameplay elements |
| Fisher et al (2019)  USA  RCT | N=119  Mothers of children aged 3-5y (low income)    **Target Population of BCTs:** Mothers | Dietary intake | **Total:** Unclear    **Taxonomy:** BCTTv1    **BCTs reported with number and label:** N | **BCT Training:** Not reported  **Selection of BCTs:** Not reported |
| Fletcher et al (2013) and Wyse et al (2014)  Australia  Cluster RCT | N=394  Parents of children aged 3-5y    **Target Population of BCTs:** Parents | Dietary intake | **Total:** Unclear    **Taxonomy:** Abraham et al 2008    **BCTs reported with number and label:** N | **BCT Training:** Not reported  **Selection of BCTs:** Not reported |
| Fulkerson et al (2015)  USA  RCT, staggered cohort | N=149  Children aged 8-12y (BMI percentile above the 50^th^ percentile) and their parents    **Target Population of BCTs:** Children and parents | Dietary intake  Sedentary behaviour | **Total:** 30    **Taxonomy:** BCTTv1    **BCTs reported with number and label:** N | **BCT Training:** Not reported  **Selection of BCTs:** Not reported |
| Gholami et al (2015)  Germany  RCT | N=155  Mothers with daughters aged 6-11y    **Target Population of BCTs:** Mothers | Dietary intake | **Total:** 4    **Taxonomy:** CALO-RE    **BCTs reported with number and label:** Y | **BCT Training:** Not reported  **Selection of BCTs:** Not reported |
| Hammersley et al (2020) (protocol)  Australia  Partially randomised preference design | N/A  Parents of children aged 2-6y    **Target Population of BCTs:** Parents | Dietary intake  Physical activity | **Total:** Not reported    **Taxonomy:** Not reported    **BCTs reported with number and label:** N | **BCT Training:** Not reported  **Selection of BCTs:** Not reported |
| Lakshman et al (2014, 2018)  UK  RCT | N= 78 (pilot) / 669 (RCT)  Mothers with infants (<14wks old)    **Target Population of BCTs:** Mothers | Infant feeding | **Total:** 13    **Taxonomy:**  Abraham et al 2008    **BCTs reported with number and label:** N | **BCT Training:** Not reported  **Selection of BCTs:**  BCTs selected were informed by the theoretical basis of the intervention (Social Cognitive Theory and Implementation Intentions) with evidence of effectiveness in changing dietary behaviours. BCTs were operationalize as intervention strategies in the intervention protocols. |
| Lin et al (2016)  China  Cluster RCT | N=1209  Parents of children aged 3-6y    **Target Population of BCTs:** Children and parents | Dietary intake | **Total:** 7    **Taxonomy:** Not reported    **BCTs reported with number and label:** N | **BCT Training:** Not reported  **Selection of BCTs:** Not reported |
| Lloyd et al (2011)  UK  Intervention development | N=119 (pilot phase 1) / 77 (pilot phase 2)  Children aged 8-11y    **Target Population of BCTs:** Children, teachers, parents/ families, school senior management | Dietary intake  Sedentary behaviour | **Total:** Unclear    **Taxonomy:** Abraham et al 2008; Michie et al 2008    **BCTs reported with number and label:** N | **BCT Training:** Not reported  **Selection of BCTs:**  Intervention mapping process to guide intervention development with feasible BCTs selected.  Suitable BCTs were identified by:   - discussions with stakeholders, and experts in behaviour change (behavioural science academics/health promotion staff) - reference to a taxonomy of behavioural change techniques - consideration of theory and practice in other school-based interventions - applying criteria for feasibility, acceptability and cost within a school setting. |
| Majumdar et al (2013)  USA  Pre-post matched pair intervention-control study | N=590  Children aged 11-13y (from schools in low income areas)    **Target Population of BCTs:** Children | Dietary intake  Physical activity  Sedentary behaviour | **Total:** 16    **Taxonomy:** Michie et al 2008    **BCTs reported with number and label:** N | **BCT Training:** Not reported  **Selection of BCTs:** Not reported |
| Marshall et al (2021)  Australia  Intervention development (cultural adaptation) | N/A  Mothers of children aged <5y (Arabic and Chinese speaking backgrounds, from socioeconomically disadvantaged areas)    **Target Population of BCTs:** Mothers | Infant feeding  Dietary intake  Physical activity  Sedentary behaviour  Sleep | **Total:** 6    **Taxonomy:** BCTTv1    **BCTs reported with number and label:** N | **BCT Training:** Not reported  **Selection of BCTs:**  Adapted existing intervention therefore did not select BCTs, instead examined the program theory of change, logic model and BCTs used. |
| Martin et al (2015)  Ireland  RCT | N=28  Students aged 8-9y    **Target Population of BCTs:** Teachers | Physical activity | **Total:** 6    **Taxonomy:** BCTTv1    **BCTs reported with number and label:** N | **BCT Training:** Not reported  **Selection of BCTs:**  The Behaviour Change Wheel, including COM-B behavioural analysis were used with BCTs selected from intervention functions mapped to specific techniques. |
| McSweeney et al (2017)  UK  Cluster RCT | N=36  Parents of children aged 3-4y    **Target Population of BCTs:** Children and parents | Dietary intake  Physical activity  Sedentary behaviour | **Total:** 12    **Taxonomy:** CALO-RE    **BCTs reported with number and label:** N | **BCT Training:** Not reported  **Selection of BCTs:**  BCTs were selected considering the targeted theoretical constructs (e.g. self-efficacy) and both individual and environmental mechanisms of action. |
| Morgan et al (2019)  Australia  RCT | N=115 fathers, 153 daughters  Girls aged 4-12yo and their fathers    **Target Population of BCTs:** Fathers and daughters | Physical activity  Sedentary behaviour | **Total:** 21    **Taxonomy:** BCTTv1, refers to additional techniques to be included BCTTv2    **BCTs reported with number and label:** N | **BCT Training:** Not reported  **Selection of BCTs:** Not reported |
| Morrison et al (2013) and Yam et al (2012)  Scotland  Exploratory RCT | N=28  Children aged 9-11y and their families (with dogs)    **Target Population of BCTs:** Children and parents | Physical activity | **Total:** 7    **Taxonomy:** Not reported    **BCTs reported with number and label:** N | **BCT Training:** Not reported  **Selection of BCTs:** Not reported |
| Phillips et al (2018)  UK  Intervention development | N/A  Pregnant women and mothers of infants (young mothers or from socioeconomically deprived areas)    **Target Population of BCTs:** Pregnant women and mothers | Infant feeding | **Total:** 22    **Taxonomy:** BCTTv1    **BCTs reported with number and label:** N | **BCT Training:** Not reported  **Selection of BCTs:**  The Behaviour Change Wheel guided intervention development process using findings of qualitative research and stakeholder consultation, and evidence from existing literature, to identify: the target behaviour to be changed, sources of this behaviour based on the COM-B model, intervention functions that could alter this behaviour, BCTs and mode of delivery for the intervention. |
| Po'e et al (2013)  USA  RCT | N=600  Parents of children aged 3-5y (high-risk)    **Target Population of BCTs:** Children and caregivers/parents | Dietary intake  Physical activity  Sleep | **Total:** 7    **Taxonomy:** BCTTv1    **BCTs reported with number and label:** N | **BCT Training:** Not reported  **Selection of BCTs:** Not reported |
| Powell et al (2016)  UK  Quasi-experimental non-equivalent group design | N=485  Children aged 7-9y    **Target Population of BCTs:** Children and teachers | Physical activity | **Total:** 3    **Taxonomy:** CALO-RE    **BCTs reported with number and label:** N | **BCT Training:** Not reported  **Selection of BCTs:**  BCTs were selected considering the SHARP Principles Model which was grounded in the Self Determination Theory, the Social Ecological Model |
| Roberts-Gray et al (2016)  USA  Cluster RCT | N=633  Parents of children aged 3-5y    **Target Population of BCTs:** Parents | Dietary intake | **Total:** 16    **Taxonomy:** Abraham et al 2008    **BCTs reported with number and label:** N | **BCT Training:** Not reported  **Selection of BCTs:**  BCTs were selected considering Social Cognitive Theory, the social-emotional-cognitive Theory of Reasoned Action, and an ecological approach that focuses on people and environments as targets for health promotion. |
| Russell et al (2018)  Australia  Quasi-experimental | N=645  Pregnant women or main caregiver of infants aged <3mths    **Target Population of BCTs:** Pregnant women and caregivers | Infant feeding | **Total:** Not reported    **Taxonomy:** BCTTv1    **BCTs reported with number and label:** N | **BCT Training:** Not reported  **Selection of BCTs:**  BCTs were selected using the Behaviour Change Wheel framework as a guide. The BCW was used to identify determinants of the target behaviours and their alternatives (less desirable behaviours) and to map these to BCTs. |
| Skouteris et al (2016)  Australia  RCT | N=250  Parents of children aged 2-4y    **Target Population of BCTs:** Parents | Dietary intake  Physical activity  Sedentary behaviour | **Total:** 6    **Taxonomy:** CALO-RE    **BCTs reported with number and label:** Y | **BCT Training:** Not reported  **Selection of BCTs:** Not reported |
| Smith et al (2018)  USA  Intervention development (adaptation) | N=350  Children aged 2-17y and their parents    **Target Population of BCTs:** Parents | Dietary intake  Physical activity | **Total:** 23    **Taxonomy:** Adapted version of BCTTv1 (JaKa et al 2017)    **BCTs reported with number and label:** Y | **BCT Training:** Not reported  **Selection of BCTs:**  BCT taxonomy was used to code content of the original Family Check-Up program and the adapted FCU4Health program, as well as rating the emphasis of the BCT on a scale from 1-5. |
| Taylor et al (2013)  UK  Intervention development | N/A  Pregnant women (BMI >25, from a multi-ethnic community)    **Target Population of BCTs:** Pregnant women | Infant feeding  Dietary intake  Physical activity | **Total:** 27    **Taxonomy:** Abraham et al 2008    **BCTs reported with number and label:** Y | **BCT Training:** Not reported  **Selection of BCTs:**  Intervention Mapping process was used to develop the intervention, including a literature review to identify theoretical determinants of behaviours, community needs assessment, develop performance objectives and select evidence-based BCTs effective in addressing specific psychological constructs. BCTs were operationalised into pragmatic and practical applications by behaviour change experts and practitioners. |
| Toomey et al (2020)  Ireland  Intervention development | N/A  Parents of infants aged 0-2y    **Target Population of BCTs:** Parents, healthcare professionals | Infant feeding  Dietary intake | **Total:** 17    **Taxonomy:** BCTTv1    **BCTs reported with number and label:** Y | **BCT Training:** Not reported  **Selection of BCTs:**  The CHErIsH team conducted five evidence syntheses and two primary qualitative studies with HCPs and parents, as well as formal and informal consultations with policy, practice, and researcher stakeholders.    The Behaviour Change Wheel framework was used to develop the intervention by an interdisciplinary research team. Evidence sources included five evidence syntheses and two primary qualitative studies with health care professionals and parents, as well as formal and informal consultations with policy, practice, and researcher stakeholders. Suitable BCTs linked with intervention functions were judged by the APEASE criteria. BCT selection included consideration of the number of barriers/enablers the BCT is relevant to, and if the BCT was identified in generally / moderately effective interventions. |
| Vaughn et al (2019) and Hennink-Kaminski et al (2018) (protocol)  USA  Intervention development / Cluster RCT | N/A  Parents of children aged <5y and childcare providers    **Target Population of BCTs:** Parents and childcare providers | Dietary intake  Physical activity | **Total:** 13    **Taxonomy:** BCTTv1    **BCTs reported with number and label:** Y | **BCT Training:** Not reported  **Selection of BCTs:**  BCTs to address behavioural determinants were selected. |

BCT: behaviour change techniques; Y: Yes; N: No; RCT: randomised controlled trial

**Supplementary Table 2:** Presence of BCTs (BCTTv1 taxonomy) in prospective studies targeting childhood obesity prevention (n=15^a^)

| **BCT number and label** | **Burton et al (2021)** | **Clarke et al (2020)** | **Draper et al (2019)** | **Espinosa-Curiel et al (2020)^b^** | **Fisher et al (2019)^c^** | **Fulkerson et al (2015)^d^** | **Marshall et al (2021)** | **Martin et al (2015)** | **Morgan et al (2019)^e^** | **Morrison et al (2013)** | **Phillips et al (2018)** | **Po'e et al (2013)** | **Smith et al (2018)^f^** | **Toomey et al (2020)** | **Vaughn et al (2019)** | **Total** |
| --- | --- | --- | --- | --- | --- | --- | --- | --- | --- | --- | --- | --- | --- | --- | --- | --- |
| **1.1. Goal setting (behaviour)** |  |  | X | X |  | X |  | X | X | X | X | X | X |  |  | 9 |
| 1.2. Problem solving |  | X | X |  | X | X | X |  |  |  | X | X | X |  | X | 9 |
| 1.3. Goal setting (outcome) | X | X |  |  |  | X |  |  |  |  | X | X |  |  |  | 5 |
| 1.4. Action planning | X |  |  |  |  | X |  | X | X | X | X |  | X |  | X | 8 |
| 1.5. Review behaviour goal(s) |  |  |  |  |  | X | X |  |  |  | X | X | X |  |  | 5 |
| 1.6. Discrepancy between current behaviour and goal |  |  |  |  |  | X |  |  |  |  |  |  |  |  |  | 1 |
| 1.7. Review outcome goal(s) |  | X |  |  |  | X |  |  |  |  | X | X |  |  |  | 4 |
| 1.9. Commitment |  |  |  |  |  | X |  |  |  |  | X |  | X |  | X | 4 |
| 2.1. Monitoring of behaviour by others without feedback |  |  |  |  |  | X |  |  |  |  |  |  |  |  |  | 1 |
| 2.2. Feedback on behaviour |  |  |  | X |  | X | X |  |  |  |  |  |  |  |  | 3 |
| **2.3. Self-monitoring of behaviour** |  |  | X |  | X | X |  | X | X | X |  | X | X |  | X | 9 |
| 2.4. Self-monitoring of outcome(s) of behaviour |  |  |  |  |  | X |  |  |  |  |  | X |  |  |  | 2 |
| 2.7. Feedback on outcome(s) of behaviour |  | X |  |  |  |  |  |  |  |  |  |  |  |  |  | 1 |
| 3.1. Social support (unspecified) |  | X |  |  |  | X | X |  |  |  | X |  | X |  |  | 5 |
| 3.2. Social support (practical) |  | X | X |  |  |  |  |  | X | X | X |  | X |  |  | 6 |
| 3.3. Social support (emotional) |  | X | X |  |  |  |  |  | X | X | X |  |  |  |  | 5 |
| **4.1. Instruction on how to perform the behaviour** | X | X |  | X |  | X | X | X | X | X | X |  | X | X | X | 12 |
| **5.1. Information about health consequences** |  | X | X | X |  | X | X |  | X | X | X |  | X | X | X | 11 |
| 5.2. Salience of consequences |  |  | X | X |  |  |  |  |  |  |  |  |  |  |  | 2 |
| 5.3. Information about social and environmental consequences |  |  | X |  |  |  |  |  | X | X | X |  |  | X | X | 6 |
| 5.4. Monitoring of emotional consequences |  |  |  |  |  | X |  |  |  | X |  |  |  |  |  | 2 |
| 5.6. Information about emotional consequences | X |  | X |  | X |  |  |  | X |  | X |  |  |  |  | 5 |
| **6.1. Demonstration of the behaviour** | X | X |  | X |  | X |  |  | X | X | X |  |  | X | X | 9 |
| 6.2. Social comparison |  |  |  | X | X |  |  |  |  |  | X |  |  |  |  | 3 |
| 7.1. Prompts/cues |  |  |  | X |  |  |  |  | X | X |  |  | X | X |  | 5 |
| 7.3 Reduce prompts/cues |  |  |  |  |  | X |  |  |  |  |  |  |  |  |  | 1 |
| 7.7 Exposure |  |  |  |  |  | X |  |  |  |  |  |  |  |  |  | 1 |
| 8.1. Behavioural practice/rehearsal |  | X |  | X |  | X |  |  |  |  |  |  | X | X | X | 6 |
| 8.2. Behaviour substitution |  |  |  | X |  | X |  | X |  |  |  |  |  |  |  | 3 |
| 8.3. Habit formation |  |  |  | X |  | X |  |  |  |  |  |  | X |  |  | 3 |
| 8.4. Habit reversal |  |  |  | X |  | X |  |  |  |  |  |  |  |  |  | 2 |
| 8.7. Graded tasks |  |  |  | X |  |  |  |  | X | X |  |  |  |  |  | 3 |
| 9.1. Credible source |  |  |  |  |  |  |  |  | X | X |  |  |  | X | X | 4 |
| 9.2. Pros and cons |  |  |  | X |  | X |  |  |  |  | X |  |  |  |  | 3 |
| 10.1. Material incentive (behaviour) |  |  |  |  |  |  |  |  | X | X |  |  | X |  |  | 3 |
| 10.4. Social reward |  |  |  | X | X |  |  |  |  |  | X |  | X |  |  | 4 |
| 10.5. Social incentive |  |  |  |  |  |  |  |  |  |  |  |  | X |  |  | 1 |
| 10.6. Non-specific incentive |  |  |  |  |  |  |  |  | X | X |  |  | X |  |  | 3 |
| 10.7. Self-incentive |  |  |  |  |  |  |  |  | X | X |  |  | X |  |  | 3 |
| 10.8. Incentive (outcome) |  |  |  |  |  | X |  |  |  |  |  |  |  |  |  | 1 |
| 10.10. Reward (outcome) |  |  | X | X |  |  |  |  |  |  |  |  |  |  |  | 2 |
| 12.1. Restructuring the physical environment |  |  | X |  | X | X |  | X |  |  |  |  | X | X |  | 6 |
| 12.2. Restructuring the social environment |  | X | X |  |  |  |  |  |  |  |  |  |  | X |  | 3 |
| 12.5. Adding objects to the environment | X |  |  |  |  |  |  |  | X | X |  |  | X | X | X | 6 |
| 13.1. Identification of self as role model |  | X |  | X |  |  |  |  | X | X |  |  | X |  | X | 6 |
| 13.2. Framing/reframing |  |  | X |  |  |  |  |  | X | X | X |  |  |  | X | 5 |
| 13.3. Incompatible beliefs |  |  |  |  |  |  |  |  |  |  | X |  |  |  |  | 1 |
| 13.5 Identity associated with changed behaviour |  |  |  |  |  | X |  |  |  |  | X |  |  |  |  | 2 |
| 14.4. Reward approximation |  |  |  | X |  |  |  |  |  |  |  |  |  |  |  | 1 |
| 14.5 Reward completion |  |  |  | X |  |  |  |  |  |  |  |  |  |  |  | 1 |
| 14.7 Reward incompatible behaviour |  |  |  | X |  |  |  |  |  |  |  |  |  |  |  | 1 |
| 15.1. Verbal persuasion about capability |  | X | X |  |  | X |  |  | X | X |  |  |  |  |  | 5 |
| 15.2. Mental rehearsal of successful performance |  | X |  |  |  |  |  |  |  |  |  |  |  |  |  | 1 |
| 15.3. Focus on past success |  |  |  |  |  | X |  |  |  |  |  |  | X |  |  | 2 |
| 15.4. Self-talk |  |  |  |  |  | X |  |  | X | X | X |  |  |  |  | 4 |
| 16.3. Vicarious consequences |  |  |  | X |  |  |  |  |  |  |  |  |  |  |  | 1 |
| **Total BCT present** | **6** | **15** | **14** | **21** | **6** | **30** | **6** | **6** | **21** | **21** | **22** | **7** | **22** | **10** | **13** |  |

^a^ Only includes studies that reported using BCTTv1. Summarises BCTs used in the intervention regardless of target population; does not list BCTs not included in any studies.

^b^ Espinosa-Curiel et al (2020) reported two additional BCTs not from the taxonomy: behaviour instigation, and mastery experiences.

^c^ Fisher et al (2019) only reported examples of BCTs included in the intervention under hierarchical clusters, therefore it is likely additional BCTs were included but not reported in the publication.

^d^ Fulkerson et al (2015) reported an additional BCTs not from the taxonomy: self-assessment of affective consequences.

^e^ Morgan et al (2019) reported an additional BCTs not from the taxonomy: increase positive emotions.

^f^ Smith et al (2018) reported an additional BCT not from the taxonomy: BCT 99.01 Information gathering.

**Supplementary Table 3:** Researcher methods to code BCT in childhood obesity prevention retrospective studies (n=23)

| **Author and year**  **Study design**  **Included studies**  **Location** | **Population** | **Target behaviours** | **Reported BCTs** | **Intervention material coded** | **Coding process** | **BCT training** | **BCTs synthesis procedure** |
| --- | --- | --- | --- | --- | --- | --- | --- |
| Anselma et al (2020) / Anselma et al (2016) (registration)  Systematic review    **Included studies:**  Total: 26  (16 RCT, 8 controlled trials)    USA, New Zealand, Canada, Northern Ireland, Mexico, Ireland, Australia, The Netherlands | Children aged 9-12y (from low socioeconomic environments)    **Target population of BCTs:** Children | Dietary intake  Physical activity  Sedentary behaviour | **Total unique:** 43 (40 from BCTTv1, + 3)  **Mean:** 6.44  **Range:** 1 to 13    **Taxonomy:** BCTTv1 and 3 additional categories added (Knowledge transfer, Community involvement, Active learning)    **BCTs reported with number and label:** Y | Published | **Coders:** 2 per study  **Independence:** Yes  **Discrepancies:** Resolved through discussion, third reviewer consulted if needed  **Agreement:** Not reported | Not reported | Narrative summary, primarily at the hierarchical cluster level |
| Azevedo et al (2019)  Systematic review    **Included studies:**  Total: 16  (12 RCT, 4 longitudinal)    UK, Europe, North America, Australia | Caregivers of children aged 0-6y    **Target population of BCTs:**  Children, parents, childcare teachers and providers | Sedentary behaviour | **Total unique:** 21^a^  **Mean:** 4.3  **Range:** 1-9    **Taxonomy:** BCTTv1    **BCTs reported with number and label:** N | Published | **Coders:** 3 in total  **Independence:** Unclear  **Discrepancies:** Resolved through discussion  **Agreement:** Not reported  **Additional info:** Coding performed by 1 reviewer and verified by two others | Not reported | Narrative summary at hierarchical cluster level of BCTTv1 and harvest plot analysis |
| Busch et al (2017)  Systematic review    **Included studies:**  Total: 11  (5 RCT, 2 controlled trials, 2 cluster RCT, interrupted time series, pre-post)    USA, China, France, Israel, Australia, Belgium Estonia,  Germany, Italy, Spain, Sweden | Children aged 4-12y    **Target population of BCTs:** Children | Sleep health | **Total unique:** 26  **Mean:** 5.27  **Range:** 3 to 15    **Taxonomy:** BCTTv1    **BCTs reported with number and label:** Y | Published | **Coders:** Not reported  **Independence:** Not reported  **Discrepancies:** Not reported  **Agreement:** Not reported | Not reported | Narrative summary |
| Farre et al (2021)  (registration)  Systematic review mixed methods synthesis with embedded stakeholder engagement    **Included studies:**  N/A | Not reported | Infant feeding | N/A    **Taxonomy:** Not reported | Not reported | **Coders:** 2 per study  **Independence:** Yes  **Discrepancies:** Resolved by discussion and involvement of a third reviewer if necessary  **Agreement:** Not reported | Not reported | Not reported |
| Hendrie et al (2017)  Systematic review    **Included studies:**  Total: 22  (8 RCT, 5 cluster RCT, 5 cohort, 2 pre-post, 1 group randomised nested cohort design, 1 randomised cross over trial)    Not reported | Children aged 2-12y    **Target population of BCTs:** Children, Parents, Teachers, Communities, Caregivers | Dietary intake | **Total unique:** 46  **Mean:** Not reported  **Range:** Not reported    **Taxonomy:** CALO-RE and 6 additional categories    **BCTs reported with number and label:** N | Published | **Coders:** 1 per study, 2 in total  **Independence:** Yes  **Discrepancies:** Inconsistencies were resolved by discussion with a third reviewer  **Agreement:** Not reported  **Additional info:** Interventions were double coded by a health psychology researcher to identify BCTs. A second coder reviewed 20% of the studies to ensure accuracy of coding. | Not reported | Narrative summary |
| Hennessy et al (2019)  Systematic review    **Included studies:**  Total: 39  (24 RCT, 13 cluster RCT, 2 quasi RCT)    USA, Australia, Finland, Brazil, Denmark, Germany, Iran, New Zealand, Sweden, The Netherlands, France  Iceland, Ireland, Italy, Republic of Belarus, Turkey, UK | Pregnant women and/or parents of children aged < 2y    **Target population of BCTs:** Pregnant women and/or parents | Infant feeding | **Total unique:** 31  **Mean:** 6.4  **Range:** 1 to 15    **Taxonomy:** BCTTv1    **BCTs reported with number and label:** Y | Published | **Coders**: 2 per study  **Independence:** Yes  **Discrepancies:** Resolved by discussion and involvement of a third party (experienced in BCT coding) if necessary  **Agreement:** Not reported  **Additional info:** Reliability of the process assessed and improved in iterative rounds of coding  Initial round sample of five trials were independently coded | 1 coder completed University College London online taxonomy training ([www.bct-taxonomy.com](http://www.bct-taxonomy.com/)) | Narrative summary |
| JaKa et al (2019)  Secondary analysis    **Included studies:**  Total: 1 (RCT, random selection of 100 participants)    USA | Parents of children aged 5-10y at risk of overweight or obesity    **Target population of BCTs:** Parents | Dietary intake  Physical activity  Sedentary behaviour | **Total unique:** 26  **Mean:** 13.9 (SD 2.8)  **Range:** Not reported    **Taxonomy:** BCTTv1 (modified to set of 26 BCTs)    **BCTs reported with number and label:** N | Unpublished materials – audio recorded and transcribed intervention sessions | **Coders:** 1 per study, 5 in total  **Independence:** Not reported  **Discrepancies:** Weekly meetings were held to discuss coding decisions and prevent coder drift.  **Agreement:** Cohen’s kappa was 0.91  **Additional info:** A randomly selected portion of sessions from N=20 participants were double coded by the lead coder to evaluate interrater reliability.  The number of unique BCTs used during the sessions was coded in five randomly selected session transcripts per participant. | All the coders completed online training (www.bct-taxonomy.com) and two additional days of study-specific training. | Exploratory analyses – general linear regressions |
| Jaka et al (2021)  Methodology study    **Included studies:**  Total: 4 (RCT)    USA | Parents of children (age unclear)    **Target population of BCTs:** Parents and children | Dietary intake  Physical Activity  Sedentary behaviour  Sleep health | **Total unique:** 78  **Mean:** 45  **Range:** 37 to 52    **Taxonomy:** BCTTv1    **BCTs reported with number and label:** Y | Unpublished | **Coders:** 4 per study, 12 in total  **Independence:** Yes, within pairs and between internal and external pairs  2 internal coders (involved in implementation of a study) coded their own intervention materials  4 external coders (2 pairs) coded the intervention materials for all studies  **Discrepancies:** Investigators and original study groups of 4 trials compared and discussed coding discrepant techniques. Final decisions made by consensus  **Agreement:** PABAK values 0.38 to 0.55  **Additional info:** Pairs met 3 times throughout coding (10%, 50% and 100% of materials) to compare independent coding and create an adjudicated version. | **All coders:** completed University College London online taxonomy training ([www.bct-taxonomy.com](http://www.bct-taxonomy.com/))  -Practiced coding an intervention design manuscript  **Internal coders only:** participated in three 1-hour discussions led by expert taxonomy tutors (University College London)  **External coders:** prior coding experience | Narrative summary |
| Johnson et al (2018)  Systematic review    **Included studies:**  Total: 18  (13 RCT, 4 cluster RCT, 1 controlled trial)    USA, Australia, others not reported | Parents of children aged 3-8y    **Target population of BCTs:** Parents | Dietary intake | **Total unique:** 23  **Mean:** 9  **Range:** 1 to 13    **BCTs from unpublished:** total additional 8    **BCTs incl. published and unpublished:**  Mean: 14.5  Range: 1 to 22    **Taxonomy:** BCTTv1    **BCTs reported with number and label:** Y | Published and unpublished | **Coders:** 2 per study (published)  **Independence:** Yes  **Discrepancies:** Discrepancies were discussed between reviewers and consensus reached.  **Agreement:** Kappa mean 0.68, range 0.49 to 1.00, PABAK mean 0.94, range 0.87 to 1.00  **Additional info:** Unpublished materials BCTs were coded by the primary reviewer, and any uncertainties discussed with a second reviewer. | Completed University College London online taxonomy training ([www.bct-taxonomy.com](http://www.bct-taxonomy.com/)) | Narrative summary |
| Johnson et al (2020) **(protocol pre-print)**, Seidler et al (2020) **(registration)**  Systematic review    **Included studies:**  N/A | Parents of infants aged <12mths    **Target population of BCTs:** Parents | Infant feeding  Dietary intake  Physical activity  Sedentary behaviour  Sleep health | N/A    **Taxonomy:** BCTTv1 | Published and unpublished | **Coders:** 2 per study  **Independence**: Yes  **Discrepancies:** Resolved by discussion and a third coder will be consulted to reach consensus if necessary  **Agreement:** Agreement of initial coding between coders will be calculated by kappa and PABAK  **Additional info:** Coded BCTs for each trial will be sent to the respective trialists to validate the coding | Completed University College London online taxonomy training ([www.bct-taxonomy.com](http://www.bct-taxonomy.com/))  Previous experience in coding BCTs (where possible) Training session prior to coding to ensure familiar with processes | Meta-regression of commonly used BCTs |
| Kassianos et al (2019)  Systematic review and meta-analysis    **Included studies:**  Total: 23  (19 RCT, 3 cluster RCT, 1 non-RCT)    USA, Denmark, South Korea, Australia, Turkey, Canada, France, Malaysia, Hong Kong, Brazil, China, Jordan | Mothers of children aged 0 to 6 weeks at baseline    **Target population of BCTs:** Mothers | Infant feeding | **Total unique:** 29  **Mean:** 4.6  **Range:** 2 to 17    **Taxonomy:** BCTTv1    **BCTs reported with number and label:** Y | Published | **Coders:** 1 per study, 3 in total  **Independence:** Yes  **Discrepancies:** Discussed and resolved in a consensus meeting of coders  **Agreement:** Moderate inter-rater reliability between coders (IRR = 0.66)  **Additional info:** Each author coded 10% of the other authors’ codes in addition to own | Completed University College London online taxonomy training ([www.bct-taxonomy.com](http://www.bct-taxonomy.com/)) | Univariate meta-regression |
| Keys et al (2020a)(registration)  Systematic review    **Included studies:**  Total: Not reported | Parents of infants aged <12mthts    **Target population of BCTs:**  Parents | Infant feeding  Sleep health | Not reported    **Taxonomy:** BCTTv1 | Published | **Coders:** Not reported  **Independence:** Not reported  **Discrepancies:** Not reported  **Agreement:** Not reported | Not reported | Narrative summary |
| Keys et al (2020b)(registration)  Systematic review    **Included studies:**  Total: Not reported | Parents of infants aged <12mthts    **Target population of BCTs:** Parents and infants | Sleep health | Not reported    **Taxonomy**: BCTTv1 | Published | **Coders:** Not reported  **Independence:** Not reported  **Discrepancies:** Not reported  **Agreement:** Not reported | Not reported | Narrative summary |
| Lewis et al 2019 (registration) and Lewis et al (2021)  Systematic review    **Included studies:**  Total: 7 (RCT)    USA, Australia, New Zealand, Malaysia | Children aged 0-5y    **Target population of BCTs:** Parents and/or children | Sedentary behaviour | **Total unique:** 24  **Mean:** Not reported  **Range:** Not reported    **Taxonomy:** BCTTv1    **BCTs reported with number and label:** Y | Published and unpublished | **Coders:** 2 per study  **Independence:** Yes  **Discrepancies:** Resolved through discussion.  **Agreement:** Interrater reliability was high (k = 0.89, 81% agreement)  **Additional info:** The intervention manuals were coded only by the first reviewer and discussed with the second reviewer. | Completed University College London online taxonomy training ([www.bct-taxonomy.com](http://www.bct-taxonomy.com/)) | Narrative summary |
| Masteller et al (2017)  Exploratory study with content analysis    **Included studies:**  Total: 3 activity trackers and associated websites    Not reported | Children    **Target population of BCTs:** Children | Physical activity | **Total unique:** 18  **Mean:** Not reported  **Range:** 8 to 14    **Taxonomy:** BCTTv1 (subset of 36 relevant BCTs)    **BCTs reported with number and label:** N | N/A  Website and tracker | **Coders:** 2 per study  **Independence:** Yes  **Discrepancies:** Resolved through discussion.  **Agreement:** Not reported | Not reported | The degree to which each BCT was incorporated into the design of the product and website was assessed using a points system. |
| Matvienko-Sikar et al (2019) and  Matvienko-Sikar et al (2016) (registration)  Systematic review    **Included studies:**  Total: 12 (8 RCTs, 3 cluster RCT, 1 non-RCT)    USA, New Zealand, Australia, UK | Pregnant women and/or parents with children aged 0-2y    **Target population of BCTs:** Parents | Infant feeding  Dietary intake | **Total unique:** 19  **Mean:** 5.1  **Range:** 1 to 11    **Taxonomy:** BCTTv1    **BCTs reported with number and label:** Y | Published | **Coders:** 2 per study, 3 in total  **Independence:** Yes  **Discrepancies:** Discussed and resolved in a consensus meeting of 3 coders, consultation with an expert in BCTs where necessary  **Agreement:** Percent agreement 80.49% | Training and experience  Training type not specified | Narrative summary |
| Mauch et al (2018)  Systematic assessment of apps    **Included studies:**  Total: 51 apps    Not reported | Families    **Target population of BCTs:** Parents/ families | Dietary intake | **Total unique:** 19  **Mean:** 3.9 (SD 1.9)  **Range:** 1 to 10    **Taxonomy:** BCTTv1    **BCTs reported with number and label:** Y | N/A  App | **Coders:** 1 per app, 2 in total  **Independence:** Yes  **Discrepancies:** Not reported  **Agreement:** The agreement between reviewers was tested in the 11 double-assessed apps using kappa (mean 0.82, range 0.66-1) and PABAK (0.97, range 0.94-1)  **Additional info:** Apps were assessed independently by a second reviewer in a random sample of 22% | Completed University College London online taxonomy training ([www.bct-taxonomy.com](http://www.bct-taxonomy.com/)) | Narrative summary |
| Seidler et al (2020)  Multi-method study    **Included studies:**  Total: 4 (RCT)    Australia, New Zealand | Pregnant women and/or parents of children aged < 6 months    **Target population of BCTs:** Parents | Infant feeding  Dietary intake  Physical activity  Sedentary behaviour  Sleep health | **Total unique:** 35  **Mean:**20.5  **Range:** 13 to 25    **BCTs from unpublished material:** Additional 63% from published materials    **Taxonomy:** BCTTv1    **BCTs reported with number and label:** Y | Published and unpublished | **Coders:** 2 per study, 4 in total  **Independence:** Yes  **Discrepancies:** Resolved through discussion  **Agreement:** PABAK ranged from 0.70 to 0.94 | Completed University College London online taxonomy training ([www.bct-taxonomy.com](http://www.bct-taxonomy.com/)) | Narrative summary |
| Smith et al (2018)  Intervention development (adaptation)    **Included studies:**  Total: 1 (randomized effectiveness-implementation hybrid trial)    USA | N=350  Children aged 2-17y and their parents    **Target Population of BCTs:** Parents | Dietary intake  Physical activity | **Total:** 23    **Taxonomy:** Adapted version of BCTTv1 (JaKa et al 2017)    **BCTs reported with number and label:** Y | Not reported | **Coders:** Not reported  **Independence:** Not reported  **Discrepancies:** Not reported  **Agreement:** Not reported  **Additional info:** Not reported | Not reported | Not reported |
| vander Kruk et al (2013)  Systematic review    **Included studies:**  Total: 24 (6 RCT, 6 pre-post, 2 cluster RCT, 1 longitudinal controlled clinical trial, 1 cluster randomized cross-over study, 3 non-randomized quasi-experimental study, 1 cross-sectional study, 1 cluster-sampled quasi-randomized crossover trial nested in a cohort,  1 longitudinal observational clinical study, 1 non-randomized clinical study, 1 randomized open trial)    Germany, Greece, The Netherlands, UK, Finland, Italy, Switzerland, Sweden, Iceland | Children aged 0-12y    **Target population of BCTs:** Parents/caregiver and/or children | Dietary intake | **Total unique:** 32  **Mean:** 12.92  **Range:** Not reported    **Taxonomy:** Adapted version of Abraham et al (2008) (Golley et al 2011)    **BCTs reported with number and label:** N | Published | **Coders:** 2 per study  **Independence:** Yes  **Discrepancies:** Discrepancies  were resolved between the reviewers by consensus and proposed to an expert in coding the BCTs  **Agreement:** Not reported  **Additional info:** An instruction manual was available on how to identify the processes and the BCTs. Pilot test of coding was conducted with 4 trials | Not reported | Narrative summary |
| Vargas-Garcia et al (2015) (protocol) and Vargas-Garcia et al (2017)  Systematic review and Meta-analysis    **Included studies:**  Total: 40 (16 RCT, 16 cluster RCT, 6 non-RCT with control group, 2 not reported)    Australia, Belgium, Brazil, Canada, Chile, Germany, Malaysia, Mexico, New Zealand, Norway, Portugal, Turkey, the Netherlands, UK, USA | Children aged 3+y, Adolescents, Adults    **Target population of BCTs:** Community | Dietary intake | **Total unique:** 26 + 2 not in taxonomy  **Mean:** 6.2 (for intervention group)  **Range:** 0 to 17    **Taxonomy:** CALO-RE    **BCTs reported with number and label:** N | Published | **Coders:** 2 per study  **Independence:** Yes  **Discrepancies:**  Discussed with a third reviewer.  **Agreement:** Not reported  **Additional info:** For the update stage, BCT coding was performed by 1 coder per study. | Not reported | Meta-analysis with BCTs as moderators |
| Webb Girard et al  (2020)  Scoping review    **Included studies:**  Total: 64 interventions    Sub‐Saharan Africa, Bangladesh, India, Central/South America, Mexico, China, Vietnam, Cambodia, Nepal, Indonesia, Pakistan, Egypt | Parents of children aged 6-24months    **Target population of BCTs:** Not specifically reported | Infant feeding  Dietary intake | **Total unique:** 28  **Median:** 6  **Range:** 2-13    **Taxonomy:** BCTTv1    **BCTs reported with number and label:** Y | Published | **Coders:** 2 per study  **Independence:** Yes  **Discrepancies:**  2 coders compared and discussed discrepancies weekly. Discrepancies were resolved in collaboration with the first author  **Agreement:** Not reported  **Additional notes:** 2 coders initially coded four articles simultaneously and reviewed each article together with the principal investigator. | Completed University College London online taxonomy training ([www.bct-taxonomy.com](http://www.bct-taxonomy.com/)) | Narrative summary |
| Zhang et al (2020) (registration)  Systematic review    **Included studies:**  Experimental or quasi-experimental study designs    Not reported | Children aged 3-12y    **Target population of BCTs:** Families | Physical activity | N/A    **Taxonomy:** Not reported | Not reported | **Coders:** Not reported  **Independence:** Not reported  **Discrepancies:** Not reported  **Agreement:** Not reported  **Additional info:** Not reported | Not reported | Realist syntheses: Context-Mechanism-Outcome configurations for individual studies |

BCT: behaviour change techniques; Y: Yes; N: No; RCT: randomised controlled trial; PABAK: prevalence-adjusted bias-adjusted kappa

**Supplementary Table 4:** Presence of BCTs (BCTTv1 taxonomy) in retrospective studies targeting childhood obesity prevention (n=15^a^)

| **BCT number and label** | **Anselma et al (2020/ 2016)^b^** | **Azevedo et al (2019)^c^** | **Busch et al (2017)^d^** | **Hennessy et al (2019)** | **JaKa et al (2019)** | **Jaka et al (2020)^e^** | **Johnson et al (2018)** | **Kassianos et al (2019)** | **Lewis et al (2019/ 2021)** | **Masteller et al 2017** | **Matvienko-Sikar et al (2019)** | **Mauch et al (2018)** | **Seidler et al (2020)** | **Smith et al (2018)^f^** | **Webb Girard et al (2020)** | **Total** |
| --- | --- | --- | --- | --- | --- | --- | --- | --- | --- | --- | --- | --- | --- | --- | --- | --- |
| **1.1. Goal setting (behaviour)** | X | X | X | X | X | X | X |  | X | X | X | X | X | X | X | 14 |
| **1.2. Problem solving** | X | X | X | X | X | X | X | X | X |  | X |  | X | X | X | 13 |
| 1.3. Goal setting (outcome) | X |  | X | X |  | X |  | X | X |  |  |  |  | X |  | 7 |
| **1.4. Action planning** | X |  | X | X | X | X | X | X | X | X | X | X | X | X | X | 14 |
| **1.5. Review behaviour goal(s)** | X |  | X | X | X | X | X | X |  | X |  |  | X |  | X | 10 |
| 1.6. Discrepancy between current behaviour and goal |  |  | X |  |  | X | X |  |  | X |  |  | X |  |  | 5 |
| 1.7. Review outcome goal(s) |  |  |  |  | X | X |  | X |  |  |  |  |  |  |  | 3 |
| 1.8. Behavioural contract | X | X |  |  |  | X |  |  |  |  |  |  |  |  |  | 3 |
| 1.9. Commitment |  |  | X |  | X | X |  | X |  | X |  |  |  | X |  | 6 |
| 2.1. Monitoring of behaviour by others without feedback | X |  |  | X |  | X |  |  |  |  | X |  |  |  | X | 5 |
| **2.2. Feedback on behaviour** | X |  | X | X |  | X | X | X | X | X |  |  | X |  | X | 10 |
| **2.3. Self-monitoring of behaviour** | X |  | X | X | X | X | X | X |  | X | X | X | X | X | X | 13 |
| 2.4. Self-monitoring of outcome(s) of behaviour |  |  | X | X |  | X |  | X | X |  | X |  |  |  |  | 6 |
| 2.5. Monitoring of outcome(s) of behaviour without feedback |  |  | X |  |  | X |  |  |  |  |  |  |  |  |  | 2 |
| 2.6. Biofeedback |  |  |  |  |  | X |  |  |  |  |  |  |  |  |  | 1 |
| 2.7. Feedback on outcome(s) of behaviour |  |  |  | X |  | X |  | X |  |  | X |  |  |  | X | 5 |
| **3.1. Social support (unspecified)** | X | X | X | X | X | X | X | X | X | X | X | X | X | X | X | 15 |
| **3.2. Social support (practical)** | X |  | X | X | X | X |  | X |  |  | X | X | X | X | X | 11 |
| 3.3. Social support (emotional) |  |  | X | X |  | X |  | X |  | X |  |  | X |  | X | 7 |
| **4.1. Instruction on how to perform the behaviour** | X | X |  | X |  | X | X | X | X | X | X | X | X | X | X | 13 |
| 4.2. Information about Antecedents |  |  |  | X |  | X | X |  |  |  |  |  | X |  |  | 4 |
| 4.3. Re-attribution |  |  | X | X |  |  |  |  |  |  |  |  |  |  |  | 2 |
| 4.4 Behavioural experiments | X |  |  |  |  |  |  |  |  |  |  |  |  |  |  | 1 |
| **5.1. Information about health consequences** | X |  | X | X | X | X | X | X | X |  | X | X | X | X | X | 13 |
| 5.2. Salience of consequences | X |  |  |  |  | X |  |  |  |  |  |  | X |  |  | 3 |
| 5.3. Information about social and environmental consequences |  |  | X | X |  | X | X | X | X |  | X | X | X |  |  | 9 |
| 5.4. Monitoring of emotional consequences |  |  |  |  |  | X |  | X |  |  |  |  |  |  |  | 2 |
| 5.6. Information about emotional consequences |  |  |  |  |  | X |  | X |  |  |  | X | X |  |  | 4 |
| **6.1. Demonstration of the behaviour** | X | X | X | X |  | X | X | X | X |  | X | X | X |  | X | 12 |
| 6.2. Social comparison | X |  | X |  |  | X | X |  | X | X |  | X | X |  | X | 9 |
| 6.3. Information about others’ approval |  |  |  |  |  | X |  |  |  |  |  |  |  |  | X | 2 |
| **7.1. Prompts/cues** | X | X |  | X | X | X | X | X |  | X | X | X | X | X | X | 13 |
| 7.4. Remove access to the reward | X |  |  |  |  | X |  |  |  |  |  |  |  |  |  | 2 |
| 7.5. Remove aversive stimulus |  |  |  |  |  | X |  | X |  |  |  |  |  |  |  | 2 |
| 7.6. Satiation |  |  |  |  |  | X |  |  |  |  |  |  |  |  |  | 1 |
| 7.8. Associative learning |  |  |  |  |  | X |  |  |  |  |  |  | X |  |  | 2 |
| **8.1. Behavioural practice/rehearsal** | X |  | X | X | X | X | X | X | X |  | X |  | X | X | X | 12 |
| 8.2. Behaviour substitution | X | X |  | X |  | X |  |  | X |  |  | X | X |  | X | 8 |
| 8.3. Habit formation | X | X | X | X | X | X |  |  |  |  | X |  |  | X | X | 9 |
| 8.4. Habit reversal | X |  |  | X |  | X |  |  |  |  |  |  | X |  |  | 4 |
| 8.6. Generalisation of target behaviour | X |  |  |  |  | X |  |  |  |  |  |  |  |  |  | 2 |
| 8.7. Graded tasks | X | X |  |  |  | X | X |  |  |  |  |  | X |  |  | 5 |
| 9.1. Credible source | X |  |  | X |  | X | X | X | X |  | X |  | X |  | X | 9 |
| 9.2. Pros and cons | X |  | X |  |  | X | X | X | X |  |  |  | X |  |  | 7 |
| 9.3. Comparative imagining of future outcomes |  |  |  |  |  | X |  |  |  |  |  |  | X |  |  | 2 |
| 10.1. Material incentive (behaviour) | X | X |  |  | X | X |  | X |  |  |  | X |  | X |  | 7 |
| 10.2. Material reward (behaviour) | X | X |  |  | X | X |  | X | X | X |  | X |  |  |  | 8 |
| 10.3. Non-specific reward | X | X |  |  |  | X | X |  | X | X |  | X |  |  |  | 7 |
| 10.4. Social reward | X |  |  | X | X | X |  |  | X | X |  |  |  | X | X | 8 |
| 10.5. Social incentive |  |  |  |  | X | X |  |  |  |  |  |  |  | X |  | 3 |
| 10.6. Non-specific incentive |  |  |  |  | X | X |  |  |  |  |  | X |  | X |  | 4 |
| 10.7. Self-incentive |  |  |  |  | X | X |  |  |  |  |  |  |  | X |  | 3 |
| 10.8. Incentive (outcome) |  | X |  |  |  | X |  |  |  |  |  |  |  |  |  | 2 |
| 10.9. Self-reward |  |  |  |  |  | X |  |  |  |  |  |  |  |  |  | 1 |
| 10.10. Reward (outcome) | X |  |  |  |  | X |  |  |  |  |  |  |  |  |  | 2 |
| 11.2. Reduce negative emotions |  | X |  |  |  | X |  | X |  |  |  |  | X |  |  | 4 |
| 11.3. Conserving mental resources |  |  |  |  |  | X |  |  |  |  |  |  | X |  |  | 2 |
| **12.1. Restructuring the physical environment** | X | X | X | X | X | X | X |  | X |  |  |  | X | X | X | 11 |
| 12.2. Restructuring the social environment | X | X | X | X |  | X |  |  |  |  |  |  | X |  | X | 7 |
| 12.3. Avoidance/reducing exposure to cues for the behaviour |  |  | X | X |  | X | X |  | X |  |  |  | X |  |  | 6 |
| 12.4. Distraction |  |  |  |  |  | X |  |  |  |  |  |  |  |  |  | 1 |
| **12.5. Adding objects to the environment** | X |  |  | X | X | X | X | X | X |  | X | X | X | X | X | 12 |
| 12.6. Body changes |  |  |  |  |  | X |  |  |  |  |  |  |  |  |  | 1 |
| 13.1. Identification of self as role model | X | X |  | X | X | X |  |  |  |  | X |  | X | X | X | 9 |
| 13.2. Framing/reframing |  |  |  |  |  | X |  |  | X |  |  |  | X |  | X | 4 |
| 13.3. Incompatible beliefs |  |  |  |  |  | X |  |  | X |  |  |  |  |  |  | 2 |
| 13.4. Valued self-identify | X |  |  |  |  | X |  |  |  |  |  |  |  |  |  | 2 |
| 14.1. Behaviour cost | X |  |  |  |  | X |  |  |  |  |  | X |  |  |  | 3 |
| 14.2. Punishment |  |  |  |  |  | X |  |  |  |  |  |  |  |  |  | 1 |
| 14.3. Remove reward |  |  |  |  |  | X |  |  |  |  |  |  |  |  |  | 1 |
| 14.4. Reward approximation |  |  |  |  |  | X |  |  |  |  |  |  |  |  |  | 1 |
| 14.6. Situation-specific reward |  |  |  |  |  | X |  |  |  | X |  |  |  |  |  | 2 |
| 14.7 Reward incompatible behaviour |  |  |  |  |  |  |  |  |  | X |  |  |  |  |  | 1 |
| 14.9. Reduce reward frequency | X |  |  |  |  | X |  |  |  |  |  |  |  |  |  | 2 |
| 15.1. Verbal persuasion about capability | X | X | X |  |  | X |  | X |  |  |  |  |  |  | X | 6 |
| 15.2. Mental rehearsal of successful performance |  |  |  |  |  | X |  |  |  |  |  |  |  |  |  | 1 |
| 15.3. Focus on past success |  |  |  |  | X | X |  |  |  | X |  |  |  | X |  | 4 |
| 15.4. Self-talk |  |  |  |  |  | X |  |  |  |  |  |  |  |  |  | 1 |
| 16.2. Imaginary reward |  |  |  |  |  | X |  |  |  |  |  |  |  |  |  | 1 |
| 16.3. Vicarious consequences |  |  |  |  |  | X |  |  |  |  |  |  |  |  |  | 1 |
| **Total BCT present** | **40** | **19** | **26** | **31** | **23** | **77** | **23** | **29** | **24** | **18** | **19** | **19** | **35** | **22** | **28** |  |

^a^ Only studies that reported BCTs using the BCTTv1 and had results available at the time of data extraction for this review are included. Farre et al. (2021), Keys et al. (2020a) and Keys et al. (2020b) were review registrations and Johnson et al. (2020) was a protocol pre-print. **Bold text** indicates commonly reported BCTs.

^b^ Anselma et al (2016/ 2020) reported an additional 3 BCTs not from the taxonomy: Community participation/involvement, Knowledge transfer, Active learning.

^c^ Azevedo et al (2019) reported a total of 21 BCTs were identified, however only 19 unique BCTs were listed in study results. ‘Other(s) monitoring with awareness’ was not directly linked to a BCT as defined in BCTTv1.

^d^ Busch et al (2017) did not report BCTs with number and label, therefore reviewers have interpreted several codes from the names reported (e.g. Persuasive argument = 15.1 Verbal persuasion about capability).

^e^ JaKa et al (2020) reported a total of 78 BCTs were identified, however only 77 unique BCTs were reported in study supplementary file.

^f^ Smith et al (2018) reported an additional not from the taxonomy: BCT 99.01 Information gathering.
